# Sentiments and Emotions for Vaccination in 2021: An International Comparison Study

**DOI:** 10.1101/2022.11.04.22281946

**Authors:** Xue-Jing Liu

**Author notes:** Correspondence concerning this article should be addressed to name, institution, postal address.

## Abstract

Comprehending how individuals feel when they discuss the vaccine is important for the immunization campaign and outbreak management during a health emergency. Online conversations provide useful information for assessing sentimental and emotional reactions to the evolutions of the pandemic and immunization program. In this study, we employ a corpus of around 58 million English tweets from users in 17 countries that discuss vaccine-related topics in the year 2021. We apply Soft Dynamic Time Warping algorithm and Time Lag Cross-Correlation approach and find that the evolutions of sentiments closely mirror the pandemic statistics. We also examine five topics connected to vaccination and discover that trust is the most predominate feeling, followed by fear, anger, and joy. Some countries reported higher emotional scores on a theme than others (people in Cuba and the United States exhibit higher levels of trust, Pakistanis and Indians express higher levels of joy, Australians and Chinese express higher levels of fear, and Japanese and British people express higher levels of anger). This study report offers a viewpoint on the public’s response to the epidemic and vaccination and aids policy-makers with preventive strategies for a future crisis.

---

The advent of the global pandemic has made vaccination one of the most critical measures for disease control. Worldwide, more than 11.31 billion doses have been administrated in 2021 (Mathieu et al., 2020). As an inevitable part of life, the vaccination has been vocalized widely on social media, including Twitter. Self-narrated texts on vaccine-related themes, such as medical instructions, queries, experience sharing, and personal thoughts, have overrun these online venues. As a result, investigations of opinion mining on the vaccine were substantially aided by the availability of a wealth of lexical data on the evolution of vaccination activities. It is valuable to harness crucial information on sentiment and emotions detected in these corpora since they have a significant impact in human activities and behavioral changes, particularly in health behavior like vaccination.

Studies revealed that the majority of tweets sentiments on Twitter were positive regarding the COVID-19 vaccination (Lyu et al., 2021). Tweets with negative tones made up about 17% and 18% of all tweets in India (Praveen et al., 2021), the united kingdom and the United States (Hussain et al., 2021) from March to November 2020. While (Liu & Liu, 2021) found that there were almost 30% of tweets with negative sentiments between November 2020 and January, 2021, and the sentiments scores fell at the end of 2020. The survey (Yousefinaghani et al., 2021) also revealed that people in the United States, Australia, India, the United Kingdom, Canada, and Ireland showed greater inclination on vaccine rejection and hesitancy then they did toward interests on vaccine in their online discourses by the end of 2020.

Several factors could also impact people’s attitudes of vaccination. One of the most influential factors affecting a vaccine’s acceptance is its quality. It describes the safety and functions of the vaccine, and includes concerns related to physiological and psychological reactions after injection. empirical evidence proved that the safety and efficacy of covid-19 vaccination significantly impacted people’s willingness to pursue a dose (Callaghan et al., 2021). Thus, people desired more information about vaccines’ side effects, clinical trials, symptoms, and other possible consequences after injection.

The individual elements included traits, attitudes, convictions, and personalities that could influence the decision to get immunized. According to studies, vaccine choices vary among persons of different sexes, ages, and ethnic backgrounds. For instance, in Slovenia, men and the elderly were more ready to receive the vaccination (Petravic et al., 2021), and in the United States, racial and ethnic minorities were found to be more reluctant for immunization (Khubchandani et al., 2021). People who lack accurate information, trustworthy knowledge of the biological mechanisms underlying vaccines (Baj-Rogowska, 2021), and assessments of decreased individual risk of infection (Ma et al., 2021) may also be hesitate to receive vaccinations.

Aspects that come into play when people engage with others are referred to as interpersonal factors. In the textual discourse about vaccinations, people frequently discuss their families, educational environment, places of employment, neighbors, and communities while deliberating whether to get vaccinated. For instance, people who were more likely to obtain the vaccine included those who worked in medicine, knew someone who had been hospitalized or died as a result of the virus (Petravic et al., 2021), and having children at home (Khubchandani et al., 2021). There were also differences in the effects of a single interpersonal element: a study in Israel indicated that unemployed people were more inclined to accept a vaccine than their employed counterparts (Dror et al., 2020), whereas a research in the United States (Malik et al., 2020) came to the opposite conclusion that working people were more likely to get vaccine.

When people lack trust in the healthcare system, public health organizations, the government, and the vaccine supplier (Larson et al., 2018), these system factors may determine how they feel about the vaccination program. According to studies that more trust in experts and institutions (Petravic et al., 2021), national vaccination strategies like immunization schedules, quantities of vaccination points, and their localization (Baj-Rogowska, 2021) would boosted people’s acceptance of immunization.

Additionally, to ensure vaccine uptake, efficient emergency risk messages are essential to ensure vaccine uptake. Misinformation may mislead the public and exacerbate the levels of vaccine hesitancy (Lewandowsky & Van Der Linden, 2021; Muric et al., 2021).

Enlightened by the aforementioned studies, we would like to complement more insights by comparing a larger dataset from more nations and adding additional evidence on the relationship between sentiments and statistics, how people’s emotions respond to hesitation factors. The objective of this study is to characterize people’s sentiments and emotions concerning Covid-19 on twitter in 17 countries.

## Methodology

We used Twitter’s API to collect tweets creased from January 1st to December 31st 2021. Each tweet included at least one of the covid-19 vaccine keywords (the full list of key works can be found in Appendix Table A). After deleting duplication, we obtained 57,999,235 tweets from 17 countries (Appendix Table B gives all of the data specifics for each country). We eliminated all URLs and punctuation marks before processing the data, tokenized every text, and deleted stop-words. We utilized VADER (Hutto & Gilbert, 2014) to quantify the sentiment of each tweet and used compound scores to denote the sentiment ratings of the tweets.

Daily data on new cases, deaths, and vaccinations for COVID-19 are collected for each nation via (Ritchie et al., 2020). Then, using the Time Lag Cross-Correlation (TLCC) method, we determined the sentimental response to each daily statistic. we measured discrepancies between the yearly sentimental evolution and each metric via soft-DTW algorithm (Cuturi & Blondel, 2017).

The correlations between sentiment and daily new cases were considered from 1st January 2021, and the association between sentiment and daily new deaths was calculated from the first day that started to reported consecutive daily statistics. the link between sentiment and new vaccination matric was measured from the first day each country began mass vaccination for its population.

To determine how Twitter users felt about different aspects of vaccination (quality, individual, interpersonal, system, and communication), we first identified keywords regarding each topic and then used a word-embedding technique to find more terms that related to these five categories. This resulted in a lexicon with a total of 7611 words (Appendix Table C). After text stemming and lemmatization, we identified the themes for each tweet and used NRCLex (Mohammad & Turney, 2013) to determine how much each emotion —trust, joy, fear, and anger—was worth for each tweet.

## Results

### Correlation of Sentiments and Statistics

All tweets were subjected to sentiment analysis, from which we calculated a sentiment score for each tweet. Positive sentiment tweets (PST) are those with scores greater than 0.05, and negative sentiment tweets (NST) are those with scores lower than -0.05. As shown in Figure 1 that, despite fluctuations, the percentages of NST were between 20% to 40% from January to June. However, the proportions have been approaching 50% since July. Several nations reported NST percentages that were greater than 50% in July, September, and December.

**Figure 1.**
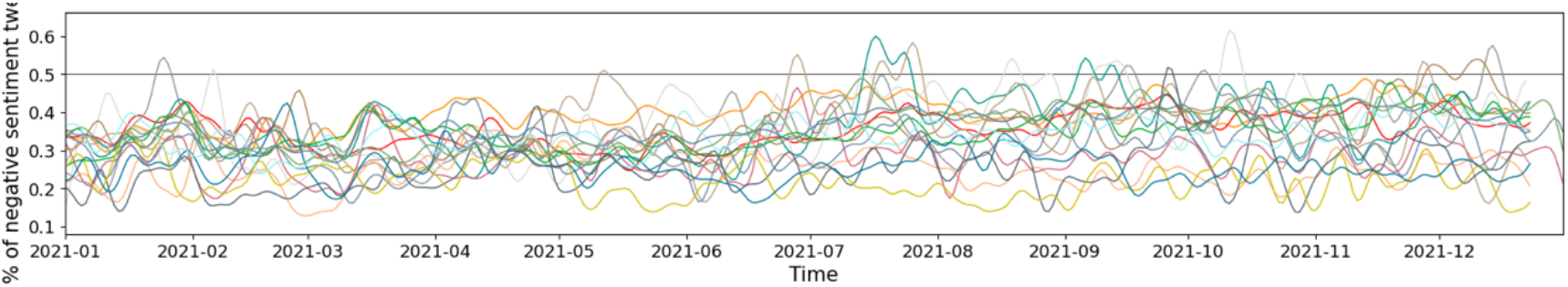
The Proportion of Negative Sentiment Tweets in 17 Countries

In Figure 2, we displayed the trends of daily new cases, new deaths, daily number of NST for each country.

**Figure 2.**
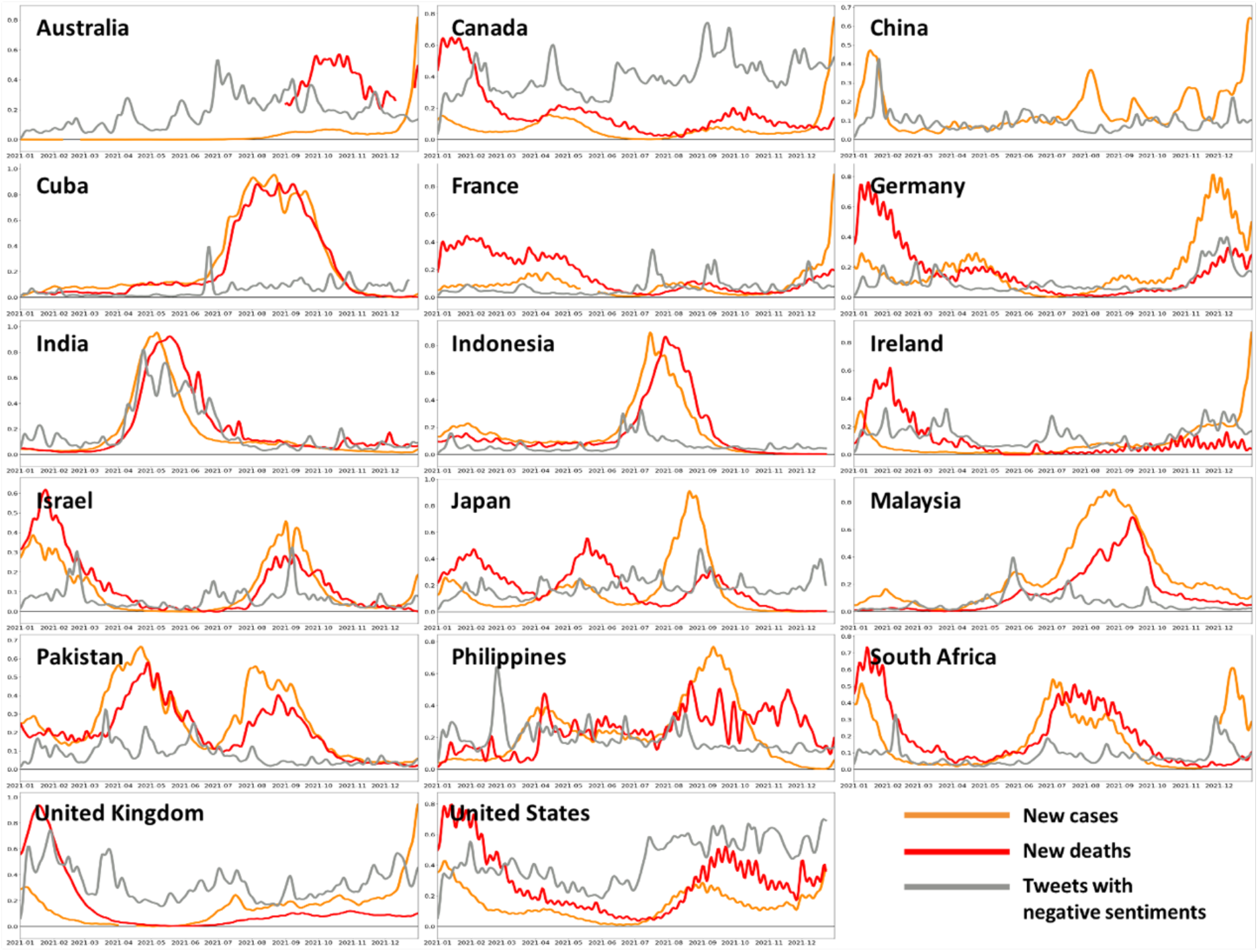
Trends of Daily New Cases, New Deaths and NST (Negative Sentiment Tweet) by Country

The discrepancies in the trends between NST and statistics differed, as shown in Table 1. It illustrated that the evolutions of negative sentiments echoed the trends of national new cases and fertilities in China, France, Germany, India and Ireland, Israel, and Japan. Meanwhile, the trends for NST in Australia, Malaysia, Pakistan, and Philippines were similar to the evolution of their fatalities.

**Table 1.** The Discrepancy Between the Trends of Negative Sentiment Tweet (NST) and Statistics Using Soft-DTW

| Country | New cases and Number of NST | New deaths and Number of NST |
| --- | --- | --- |
| Australia | 22.602 | 12.880 |
| Canada | 72.056 | 56.611 |
| China | 9.253 | / |
| Cuba | 66.358 | 56.294 |
| France | 6.342 | 17.466 |
| Germany | 13.055 | 18.463 |
| India | 6.515 | 6.120 |
| Indonesia | 20.261 | 22.782 |
| Ireland | 8.576 | 9.113 |
| Israel | 8.744 | 12.828 |
| Japan | 20.567 | 16.303 |
| Malaysia | 55.645 | 19.063 |
| Pakistan | 35.212 | 17.950 |
| Philippines | 23.899 | 14.299 |
| South Africa | 17.255 | 26.000 |
| United Kingdom | 35.852 | 39.179 |
| United States | 56.403 | 36.076 |

We computed daily sentimental scores for each country by taking the average score for users in it. Figure 3 depicts the patterns in new vaccines and daily sentiment scores. Several countries (Canada, Germany, France, Ireland, Israel, and the United States) launched national mass vaccination programs at the end of 2020, followed by nine countries which started in the early month of 2021, and Cuba and Pakistan started in June, and South Africa started in September 2021. The majority of countries reported one or two plateau periods of daily new vaccination, the first was usually between June and September, lasting one or two months, and the second was around December. Significantly, sentiment scores fluctuated across all countries, revealing two major trends: seven countries (Indonesia, India, Japan, Malaysia, Pakistan, Philippines, and South Africa) reported almost positive sentiments throughout the year, while sentiment scores in the remaining ten countries bounced around neutral, with scores in the second half of 2021 lower than those in the first half.

**Figure 3.**
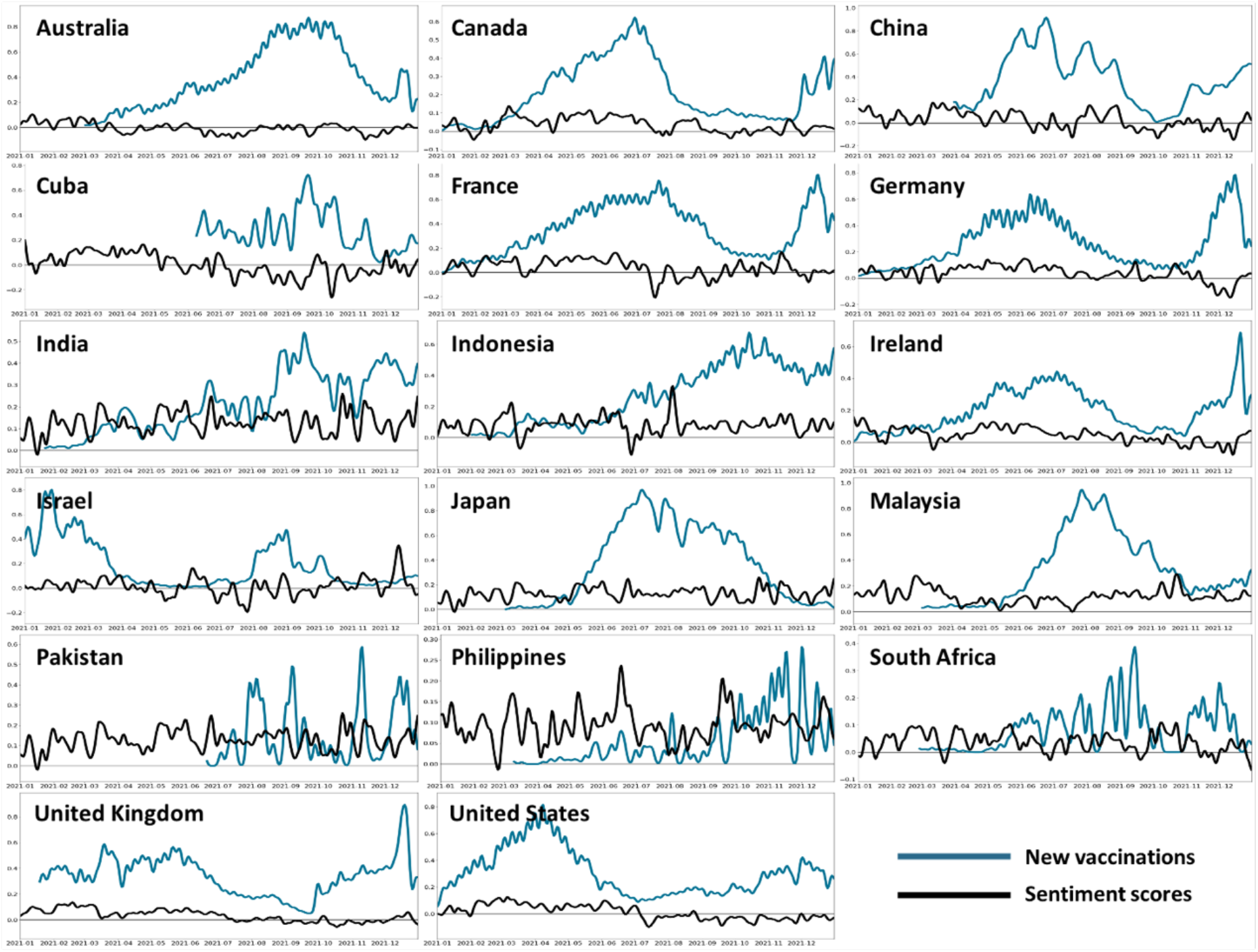
Trends of Daily New Vaccinations and Average Sentiment Scores by Country

Table 2 displays the results of the association between sentiment and daily statistics. We classified a quick response as 0 to 5 lay-days, a medium response as 6 to 10 lay-days, and a sluggish response as 11 to 15 lay-days.

**Table 2.**
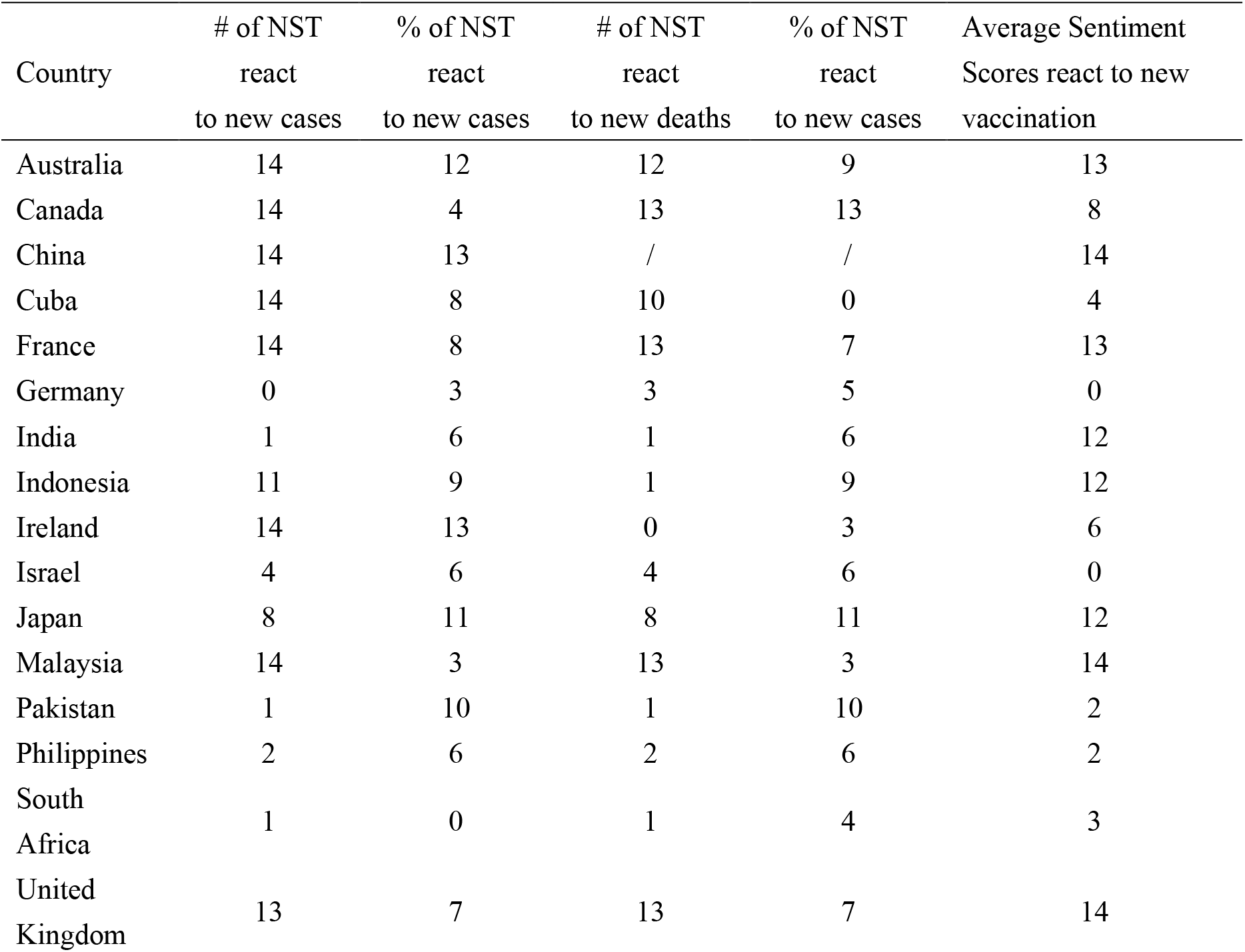
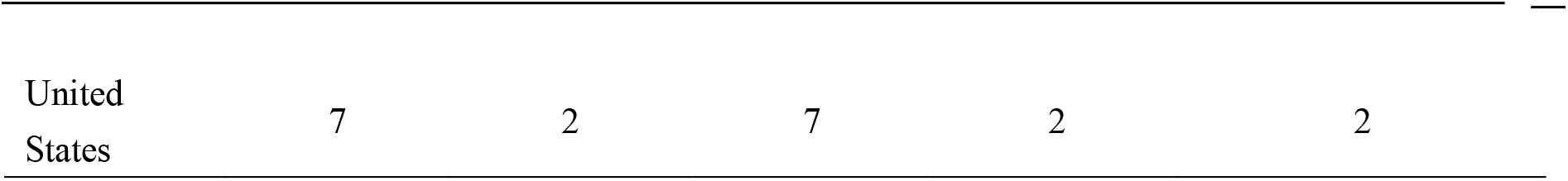
Estimated Lag Day (LagD) that Negative Sentiments Tweets (NST) and Average Sentiment Scores React to Daily New Cases, Deaths and Vaccinations

| Country | # of NST<br>react<br>to new cases | % of NST<br>react<br>to new cases | # of NST<br>react<br>to new deaths | % of NST<br>react<br>to new cases | Average Sentiment<br>Scores react to new<br>vaccination |
| --- | --- | --- | --- | --- | --- |
| Australia | 14 | 12 | 12 | 9 | 13 |
| Canada | 14 | 4 | 13 | 13 | 8 |
| China | 14 | 13 | / | / | 14 |
| Cuba | 14 | 8 | 10 | 0 | 4 |
| France | 14 | 8 | 13 | 7 | 13 |
| Germany | 0 | 3 | 3 | 5 | 0 |
| India | 1 | 6 | 1 | 6 | 12 |
| Indonesia | 11 | 9 | 1 | 9 | 12 |
| Ireland | 14 | 13 | 0 | 3 | 6 |
| Israel | 4 | 6 | 4 | 6 | 0 |
| Japan | 8 | 11 | 8 | 11 | 12 |
| Malaysia | 14 | 3 | 13 | 3 | 14 |
| Pakistan | 1 | 10 | 1 | 10 | 2 |
| Philippines | 2 | 6 | 2 | 6 | 2 |
| South<br>Africa | 1 | 0 | 1 | 4 | 3 |
| United<br>Kingdom | 13 | 7 | 13 | 7 | 14 |
| United States | 7 | 2 | 7 | 2 | 2 |

Notably, people in Germany, India, Israel, Pakistan, Philippines, and South Africa responded quickly to increased new cases by posting more NST in the next 0 to 5 days, and the percentages of those negative posts also increased in Canada, Germany, Malaysia, South Africa, and the United States in the following few days regarding to statistics changing. In comparison to new cases, users in more countries (Germany, India, Indonesia, Ireland, Israel, Pakistan, Philippines, and South Africa) post NST online, and the percentage of NST in six countries (Cuba, Germany, Ireland, Malaysia, South Africa, and the United States) respond to new death trends within five days. People’s average sentiment in Germany, Cuba, Israel, Pakistan, Philippines, South Africa, and the United States were more sensitive to daily new vaccination (LagD=0 to 4).

### Emotions for Vaccine-related Factors

For each tweet, we detected four emotions: trust, joy, anger, and fear, and estimated the average degree of them for each vaccine-related factor. To understand how people’s emotions relating to each parameter, we presented the emotions of tweets in 2021 based on five major factors.

Among these four emotions, trust played the most prominent role, with an average score for a topic ranging from 0.101 to 0.154, followed by fear (0.062-0.093), anger (0.032-0.052), and joy (0.032-0.047). Figure 4-8 depicts how people in various countries vocalized their feelings about vaccine topic.

**Figure 4.**
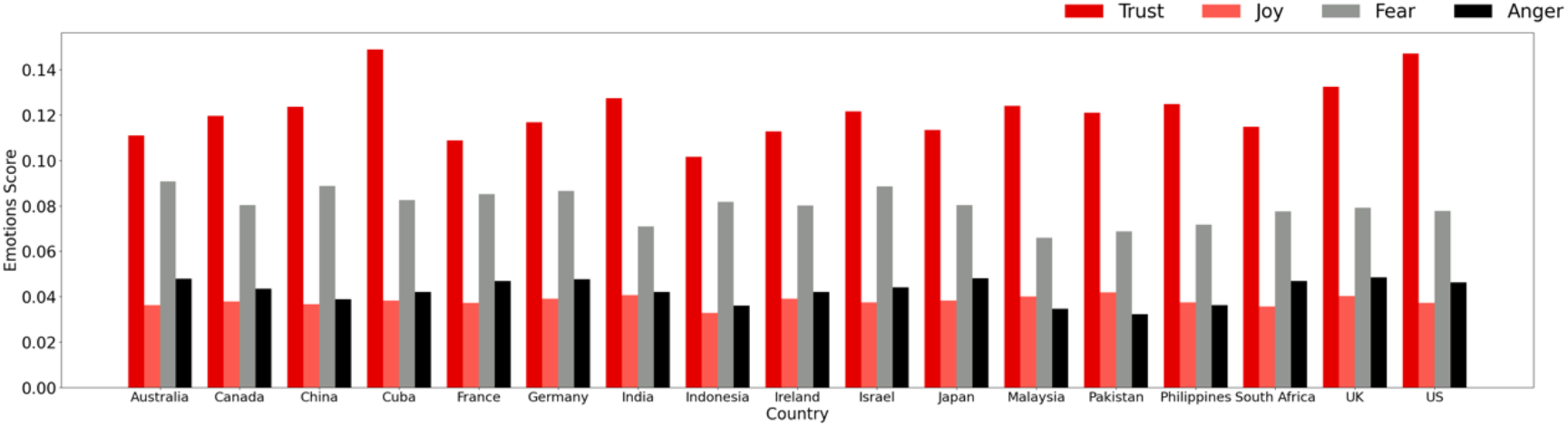
Emotions on the Quality Factors of Vaccination

**Figure 5.**
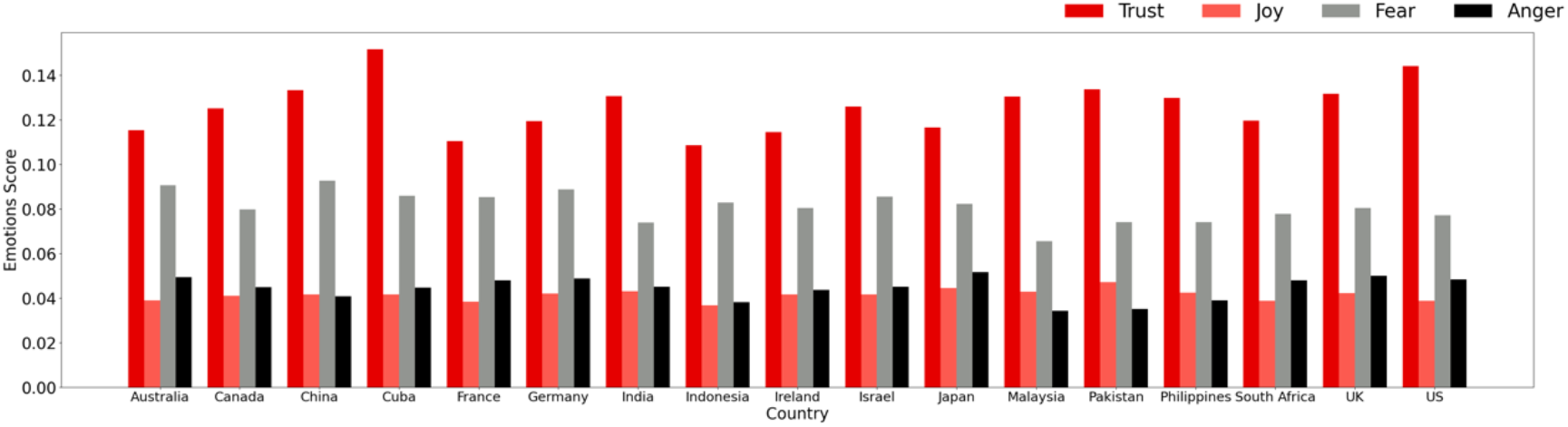
Emotions on the Individual Factors of Vaccination

**Figure 6.**
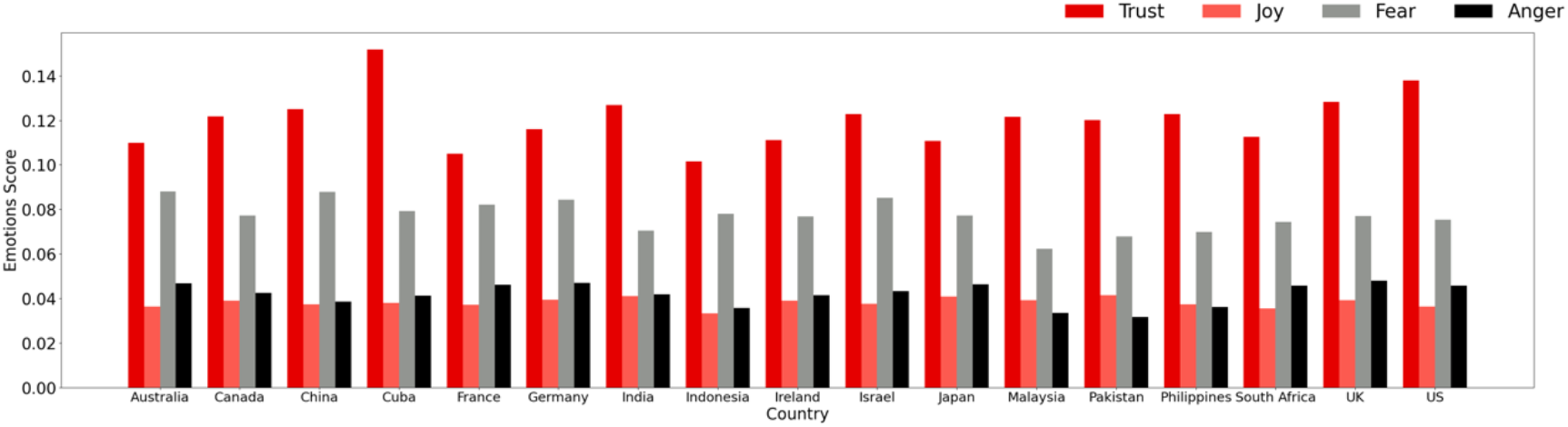
Emotions on the Interpersonal Factors of Vaccination

**Figure 7.**
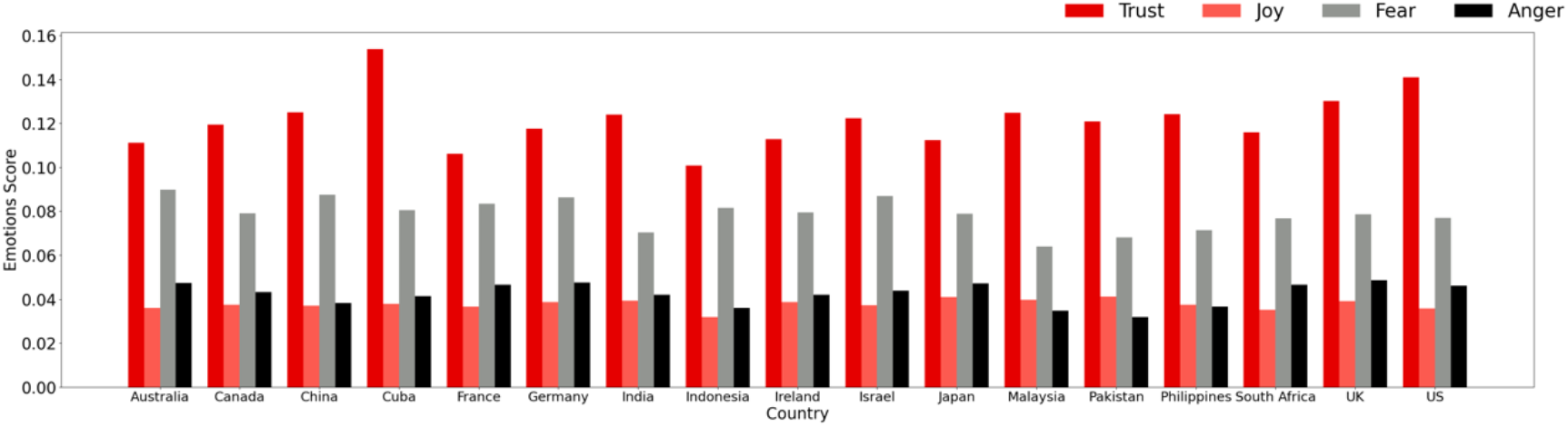
Emotions on the System Factors of Vaccination

**Figure 8.**
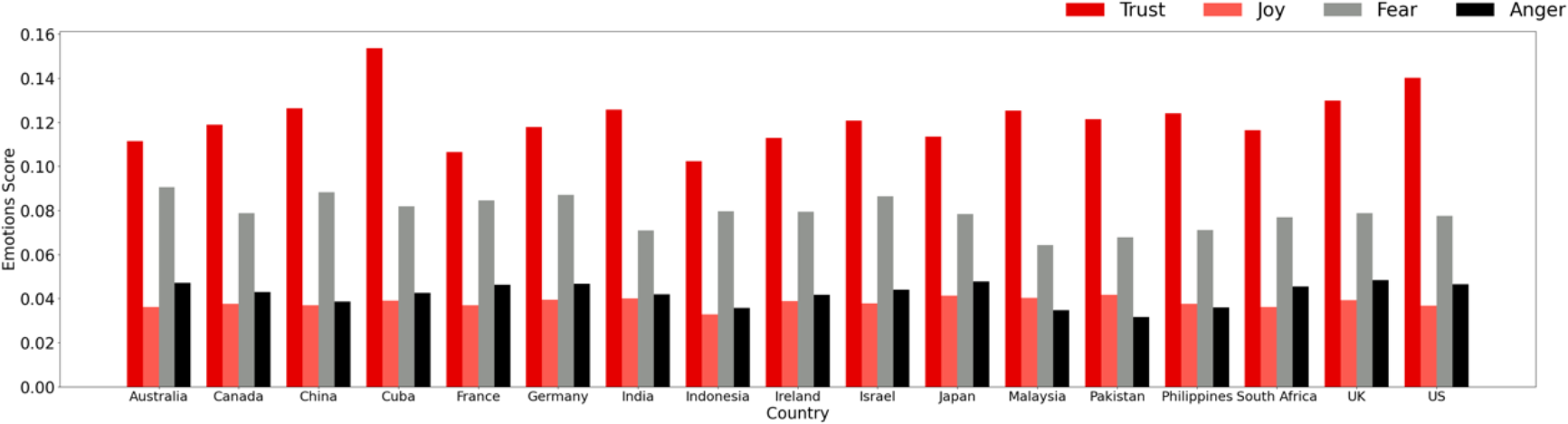
Emotions on the Communication Factors of Vaccination

As in Figure 4, Cubans had the greatest average level of trust regarding vaccine’s quality, followed by the United States, the United Kingdom, and India. People in Pakistan, India, and the United Kingdom were more likely to be joyous. On the contrary, we discovered that users in Australia, China, and Israel expressed higher levels of fear, and users in the United Kingdom, Japan, and Australia expressed more rage regarding the safety and efficiency of the vaccine products.

Showed in figure 5, people in Cuba, the United States, and Pakistan shown greater trust in the context of vaccination. At the same time, Pakistan, Japan, and India showed higher levels of happiness. While China, Australia, and Germany reported higher levels of fear in their texts. Japan, the United Kingdom, and Australia reported higher levels of anger as a result of personal traits.

When mentioned interpersonal factors, people in Cuba, the United States, and the United Kingdom expressed more trust and people in Pakistan, India, and Japan expressed more delight. However, persons in Australia, China, and Israel exhibited more fear and people in the United Kingdom, Germany, and Australia showed more anger in their texts when they were concerned about their families, friends, and communities.

People in Cuba, the United States, and the United Kingdom expressed the most trust in the system conducting the program, and people in Pakistan, Japan, and Malaysia expressed more joyful emotions. In contrast, Australia, China, and Israel showed more fear on the system and the United Kingdom, Germany, and Australia are angrier with it.

When discussing vaccination communication, Cuba, the United States, and the United Kingdom displayed greater levels of trust. The media’s coverage of vaccine information was met with greater levels of joyfulness in Pakistan, India, and the United Kingdom. Additionally, tweets from the United Kingdom, Japan, and Australia raised greater fear about the communication program while comments from Australia, China, and Israel featured more anger.

## Discussion

This study outlined the characteristics of the sentiment and emotions toward vaccination in 17 countries. We evaluated the correlation between the evolutions of the sentiments and pandemic statistics to reveal their lay-time relationship and holistic resemblance. Since June, the sentiments for online discourse have been worse as seen by an increase in the percentage of tweets with negative sentiment and a decline in the average sentiment scores. The evolution of people’s online sentiment expression was influenced by daily new cases, death, and vaccination, and their sentiments reacted to these figures with changes in time and discrepancies in trends. Finally, we looked at the emotions people had about various vaccine-related aspects, and these feelings were consistent within one country but varied across nations.

The findings in this study suggested that the public’s sentiment towards vaccination deteriorated after vaccine announcement. This may due to dissatisfactions on national extended vaccination program. Israel, the United Kingdom and the united states had finished their first waves of mass vaccination on July, other countries were in the middle of mass vaccination, and their sentiment scores decreased. One of the potential explanations for this might be the shortage of the vaccine and discussion of the priority for vaccination (WHO, 2021). At the same time, the merge of some more contagious variant, like Delta, which bring surged new cases and fertilities in some countries and reduced the efficacy of current vaccine (“WHO releases global COVID-19 vaccination strategy update to reach unprotected,” 2022). These may cause more negative sentiments toward the technology and confidence on vaccination program.

Another reason for this downward trend may be a consequence of some bad experience of vaccine service. undesirable service features, like multiple doses, long waiting time, and vaccine facilities (Eshun-Wilson et al., 2021) are barriers for pleasant experience, and messy procedure, busy phone-line, crashed website, unfriendly administration (Niu, Liu, Kato, Shinohara, et al., 2022) could trigger adverse sentiment.

The results of discrepancies between statistics and the number of posts with negative sentiments demonstrated that the evolutions of daily cases and fertilities can be good predictors on feeling of online discourse. While the discrepancies in the results of Canada, Cuba, the United Kingdom, and the united states were larger than that in other countries, because the number of tweets with negative sentiments were consistently low in Cuba and were consistently high in Canada, the united kingdom and the united states all year round no matter of the changes of statistics.

We also estimated the how many days people’s sentiments respond to the statistics and found that the number or the proportion of tweets with negative sentiment in most countries reacted promptly to the statistics and the average sentiment scores in several countries responded to vaccination progress quickly.

These findings provide supports for future vaccine campaign and communication. With the help of precise correlation predictions between statistics and sentiment, policy-makers can monitor and predict the development of people’s feeling and adjust strategies accordingly.

The findings about people’s emotions suggested that the Cubans, Americans, and British people expressed higher trust in their discourses on topics about vaccination. Pakistan and Indian people used joyous tones when they talked about the vaccination, the Japanese delivered their joy on individual, interpersonal and system factors, and British people felt more joyous about the quality and communication factors of the vaccine.

On the contrary, Australian and Chinese reported higher fear scores on most topics, Israeli fear of quality, interpersonal and system factors, German fear of interpersonal factors, and British and Japanese fear of communication factors. Higher anger scores were detected among the Australians and British on most vaccine factors, German reported higher levels of anger on interpersonal and system factors, and Japanese reported higher anger scores on quality and individual topics.

Cuba’s strategy for disease prevention is to develop its own vaccines, and by march 2021, five candidates had been created (Taylor, 2021). The country implemented a comprehensive immunization program, both its speed and extent were among the highest in the globe (Kazemi et al., 2022). And as of January 2021, 87% of the population was fully vaccinated (Aguilar-Guerra et al., 2021), which made it achieve the third highest vaccination rate in the world (Aguilar-Guerra & Gorry, 2021). At the beginning of the outbreak in Cuba, the Continuous Assessment and Risk Evaluation (CARE) system dispatched medical teams to conduct systematic door-to-door medical surveys and reacted immediately for case identification, disease prevention, and vaccination promotion (Aguilar-Guerra & Gorry, 2021). Additionally, Cuba has advances in biotech research and novel vaccine development, giving people confidence in the vaccinations’ outcomes.

Americans also reported their trust in vaccination when they talked about all the related factors. According to research (Pogue et al., 2020), one of the key indicators of vaccine acceptability was the belief that COVID-19 was a severe issue for the entire nation and the vaccination was necessary to reduce the disease’s future economic, social, and public consequences. Moreover, having faith in medical professionals, authorities, communication programs, and easy access to the vaccine (Salmon et al., 2021) improved Americans’ willing for get vaccinated. Several studies revealed that 60% to 70% of Americans supported vaccinations and roughly 84% of parents would agree to vaccinate their children if necessary (Funk & Tyson, 2020; Pogue et al., 2020; Yılmaz & Sahin, 2021). With the aid of some promotion initiatives, such as the Operation Warp Speed program (Howard & Wright, 2021), the United States had addressed structural barriers of cost, convenience, and supply (Fisk, 2021) from the national rollout. People’s trust grew as a result of timely released information by the government (Dooling et al., 2021), and by February 2022, 64% of the population had received fully vaccinations (Ritchie et al., 2020).

The implementation in Great Britain began earlier than in other nations, and the efficacy of the immunization was validated by randomized controlled studies (Voysey et al., 2021) and real-world data from vaccine recipients(Vasileiou et al., 2021). The UK made vaccinations easily accessible by providing a wide range of facilities, including hospitals, GP-led clinics, and neighborhood pharmacy sites. In addition, it released papers, used electronic GP records (Majeed et al., 2021), and tracked vaccination uptake, safety, and effectiveness (UK, 2020). These efforts provided recipients with up-to-date messages and improved transparency of the program. And it reported a lower overall rates of vaccine hesitancy than many other nations (Majeed et al., 2022), which may remove barriers brought on by individual views or historical causes. By March 2022, a booster had been delivered to 70% of those over 12-year-old in the United Kingdom (The Visual and Data Journalism Team, 2022). When people discussed vaccine-related subjects on Twitter, these elements contributed to a sense of trust and joyous. Meanwhile, a higher level of anger was reported by British people on Twitter. This negative emotion may ignite by structural racism and the inequities for ethnic minorities (Hussain et al., 2022), those groups had the lowest vaccination rate compared with white people (Nellums et al., 2022).

Socio-cultural (Ali et al., 2021), religious convictions, mistrust of the government (Khan et al., 2015; Shah et al., 2019), and false narratives (Khan et al., 2020) have had an impact on the public’s choice to be immunized in Pakistan. To improve its vaccination rate, the government made arrangements for people who live in remote areas to receive vaccines, such as the Rural Development Foundation to assist district health departments in reaching people in villages (Norwegian Church Aid, 2022), and using Expanded Immunization Program staff in far-flung areas for risk communication and door-to-door vaccination services (Basharat, 2022). Additionally, the research from 2022 revealed that 77% of participants felt that everyone should receive the immunizations provided by the government, and more than 80% of respondents were willing to accept COVID-19 vaccination (Kumar et al., 2022).

As the world’s largest manufacturer and worldwide distributor of vaccines (Lahariya, 2014; Sharun & Dhama, 2021), India has sufficient manufacturing capability and a robust strategy for vaccine storage and distribution throughout the pandemic. The government also launched training programs for immunization officers, cold-chain officers, IEC (Information, education, and communication) officials, medical workers, as well as midwives and auxiliary nurse in rural areas (Kumar et al., 2021), the vaccine acceptance rate in India is approximately 74%. (Chakraborty et al., 2021). And the Indian Minister of Health declared the end of the epidemic on early March 2021.

In comparison to other nations, Australia had a low rate of COVID-19 infection and fatalities. Between August 2020 and January 2021, there was a significant rise in vaccine hesitancy, and individuals who had less trust in the government, hospitals, or healthcare system and a lower risk perception of the disease were more likely to refuse vaccination (Biddle et al., 2021). Additionally, on April 8, thrombosis with thrombocytopenia syndrome was found to be associated with the vaccine, which led to vaccine reluctance and delay (Attwell et al., 2021). Due to their negative experiences with the media, the quality of the products, changes in policy, and the accessibility of the vaccines, two-thirds of individuals were dissatisfied with the deployment of the vaccines by April 2021 (Attwood, 2022). What’s more, according to a model (Zachreson et al., 2021), even though 82% of the population has received a vaccination, herd immunity has not been achieved in Australia due to the presence of densely clustered, networked groups. These variables may reduce public trust in the immunization program and raise levels of fear and anger expressed online.

In China, vaccination opinions have evolved. Over 90% of vaccine acceptance was reported in a research from August 2020 (Wang et al., 2020), while another study reported a 67% acceptance rate in May 2021 (Wang et al., 2021) when the vaccine is available. Because its novelty, people have less faith in the vaccination product themselves than they do in the government and the vaccine delivery system, 10% of people in the study reported trust in vaccine while denial to be vaccinated (Wang et al., 2021), which indicated low-risk susceptibility and severity perceptions of the disease in 2021. People were also concerned about the vaccine’s safety and procedure. Compared to primary health care centers, they preferred to be vaccinated in big and municipal hospitals (Leng et al., 2021). The fear feelings may be a result of the events stated above, findings from current study was consistent with another study that assessed emotions using a corpus acquired from a Chinese social media platform Weibo (Dang & Li, 2022). It explained that fear emotion may occur as a result of adverse effects and probable punishment by communities and units if people refuse to be vaccinated.

The qualitative, interpersonal, and system factors were reported to be more feared in Israel. Israel had started a vigorous vaccination campaign and was the world’s leader in immunization rollout (Kershner), but it had disparity in the program. The uptake had been much lower in the population with lower socioeconomic status, Arab population, and young women (Saban et al., 2021). Parents are reluctant to vaccinate their children due to side effects, novelty, and effectiveness of the vaccines (Savitsky et al., 2022). At the same time, the Green Pass policy (Kamin-Friedman & Peled Raz, 2021) restricted the movement of unvaccinated people was seen as inconsistent with fostering trust. That could account for why there was more dread voiced online.

According to a survey, people would get angry if the vaccination were made mandatory in Germany (Sprengholz et al., 2021), and more than half of Germans expressed opposition to such a program (Graeber et al., 2021). People also objected to utilizing “immunity cards” (Brown et al., 2021) to enter public spaces. Since July 2021, there have been an increasing number of missed vaccination appointments because some individuals think that one dose is sufficient for avoiding the disease and they did not want the vaccine to interfere with their vacation (Reuters, 2021). Thus, they showed their anger online.

Japan has over 26% of its population above the age of 65 (Yui et al., 2017), and a lower vaccine confidence indexes round the globe (De Figueiredo et al., 2020), these put more challenges for vaccination coverage. Research revealed that the acceptance rate was 60%, and that it was lower for women, those in their 20s and 50s, and those who had low income levels (Machida et al., 2021; Yoda & Katsuyama, 2021). Nevertheless, when the vaccination became available in February 2021, the percentage dropped to 47% (Kadoya et al., 2021). Lower confidence in vaccine product (Niu, Liu, Kato, Aoyama, et al., 2022) might compromise how strongly people feel about herd immunity online.

The current study has certain limitations. First of all, it is challenging to link a person’s feelings to their actual behaviors, especially when doing so at the individual level. Therefore, in order to estimate behavior and then establish a causal link, more information from users’ tweets must be taken into account.

Furthermore, because the samples were primarily made up of young, educated, urban, and digitally capable individuals (Danabal et al., 2021; Thiagarajan, 2021; Umakanthan et al., 2021), research employing online textual data may overstate attitudes and feelings about vaccinations. The conclusion should be supported by other forms of research.

Additionally, we exclusively used English-language tweets and disregarded voices in other languages. It is nevertheless important to carry out international comparative research using various languages, even though certain outcomes of the current study were compatible with the studies using their native language corpus.

Last but not least, it is challengeable to assign more accurate sentiment and mood scores to different topics discussed in a single tweet. Future research should take into account techniques in natural language processing for topic separation and aspect identification.

## Data Availability

All data produced in the present study are available upon reasonable request to the authors

