## Appendix Materials for "Sentiments and Emotions for Vaccination in 2021: An International Comparison Study"

### Table A

*Keywords for Collecting Tweets*

| |
|---|
| vaccine |
| vaccination |
| vaccinate |
| pfizercovidvaccine |
| pfizervaccine |
| covid pfizer |
| pfizer |
| moderna |
| covid biontech |
| covid_19 biontech |
| azvaccine |
| coronavaccine |
| coronavirusvaccine |
| covax |
| covid19vaccine |
| covidvaccine |
| gavi |
| glyphosate |
| mrna |
| nvic |
| oxfordvaccine |
| pharmagreed |
| rna |
| vax |
| johnson&johnson |
| sputnik |
| covaxin |
| sinovac |

### Table B

*Tweets Number from Each Country*

| Country | Number of tweets |
|---|---|
| Australia | 3,537,572 |

| | |
|---|---|
| Canada | 4,355,409 |
| China | 485,819 |
| Cuba | 445,557 |
| France | 556,550 |
| Germany | 511,121 |
| India | 7,838,045 |
| Indonesia | 191,960 |
| Ireland | 630,020 |
| Israel | 453,491 |
| Japan | 338,193 |
| Malaysia | 737,675 |
| Pakistan | 439,320 |
| Philippines | 541,410 |
| South Africa | 1,365,824 |
| United States | 26,696,951 |
| United Kingdom | 8,874,318 |

**Table C**

*Keywords for Five Vaccine-related Factors*

| Keywords for quality factor |
|---|
| consequence effect outcome impact aftermath punishment cause upshot consequently repercussion resultant reward aftereffect byproduct corollary risk justification reverberation resulting issue inevitable contributory contributing precipitated proximate response implication reaction ramification symptom immunity benchmark fever sore cough shortness breathing difficulty breath throat fatigue tiredness heart respiratory physical chest fast-beating neurological pounding brain headache sleep dizziness pins-and-needles smell depression digestive palpitation diarrhea stomach taste lightheadedness muscle rash menstrual anxiety safe comfortable secure safety dependable harmless protect strongbox protected dangerous reliable prophylactic rubber condom safest refuge healthy sane unsafe unhurt securely riskless unadventurous responsibly stable appropriate effective convenient foolproof fireproof trustworthy pain achy chill node nausea swollen unwell redness mental swelling vomiting lymph mobility diarrhoea vulnerable hazardous risky insecure precarious unhealthy unsecured harmful uncomfortable unsanitary unsuitable substandard unfit inadequate unsound unacceptable unhygienic unusable deplorable unreliable overcrowded inhumane treacherous perilous dicey suicidal bad desperate dodgy insidious parlous breakneck shaky chancy unreasonable outdated unprotected chanceful touch-and-go self-destructive unguaranteed defective unstable probability hazard danger gamble |

adventure peril jeopardy threat venture assay moral harm exposure potential possibility prevention stress burden riskier uncertainty potentially benefit susceptible counterparty risked imminent disease sign diagnosis syndrome chronic inflammation symptomatic paresthesia indication medicine sickness malady illness manifestation prognosis ailment hypertension evidence abnormality patient ague dyspnoea rhinorrhea cyanosis prodrome hemoptysis thrombocytosis haematuria treatment abdominal problem health wellness welfare well-being obesity nutrition hygiene healthcare fitness medicare nursing wellbeing epidemiology insurance sanitation damage tobacco infectious condition aid healthier shield stamen immune resistant virus microorganism cancer infection unaffected phagocytosis bacteria vaccination immunology autoimmune bacterial nervous antibody tissue louis unsusceptible lymphocyte insusceptible pathogen immunodeficiency cell phagocyte cholera impervious hiv organ gene nerve lung skin proof clinical die infect kill inllness servere mrna experiment dna fetal allergic fda adverse serious hospitalized suffering paralysis violence experience endure tolerate hurt worsen ache bear stand abide pleasure brook twinge grieve experiencing ail debilitating anguish torment afflict recover inflict hardship sustain digest stick agonize hunger collapse decline feel painful sorrow misery distressfully affect penance succumb survive suffer dying undergo spared recovering inflicting valence recuperating afflicting stricken debilitated crippling death battling affliction reeling succumbing succumbed struggling lingering distress sustained hurting ravaged evil devastating recuperated torture horrendous heartache worsening malnourishment agony inflicts ravaging crippled overcoming sufferer recovers wracked bedridden ravage suffered discomfort sustaining trauma hemorrhaging happiness plaguing convalescing diagnosed exploitation recuperate severe befallen exhaustion died killing aching struggle cruelty worsened poverty abuse loss aggravating suffers healed oppression horrific sick depressed undergoing dead accurate consistent credible trusty authentic certain authoritative precise efficient tested useful viable trusted solid inexpensive timely knowledgeable durable undeviating tried sophisticated flexible reputable adequate dependability accuracy quality time-tested competitively predictable scaleable toxic destructive detrimental abusive injurious noxious hurtful damaging deleterious pernicious prejudicial nocent mischievous malign ill traumatic subtle negative catastrophic counterproductive undesirable poisonous hateful immoral wrong corrosive deadly degrading offensive addictive slanderous disadvantageous useless defamatory violent calumnious libelous libellous ruinous beneficial problematic unfavourable denigrating fatal denigratory counter annoying wounding stabbing necessity inevitability requirement urgency essential indispensable requisite importance prerequisite desideratum convenience imperative utility unneeded obligation consideration contingency requisiteness rationale desperation unnecessary neediness absolute necessitates sufficiency survival paramount usefulness efficacy utmost preferable exigency incentive grocery staple effectiveness feasible indispensible dispensable desirability greed priority unpleasant terrible wrenching excruciating agonizing afflictive awful poignant harrowing torturous bitter frustrating painfully embarrassing painless dreadful opioid horrible irritating sensitive unspeakable troublesome abominable atrocious agonized tender biting fibromyalgia itchy galled chafed torturing scary racking awkward stressful phantom saddle-sore torturesome sorrowful agonising analgesia complicated unpleasantness expensive tight grievous gut slow tragic brutal weird intense traumatizing humbling frightening burning disheartening impossible invasive boring terrifying stiff horrifying jarring rough injury cathartic unsettling miserable cystitis irregular depressing thalamus demoralizing distressing gross dispiriting emotionally inflamed numb shameful nociceptors exhausting bothersome toothache bloody red hilarious cringe bittersweet unavoidable humiliating messy inconvenient challenging numbing lamentable gruesome verification validation substantiation prove confirmation demonstration authentication verify substantiate demonstrate proofread bulletproof weatherproof cogent exercised ordeal irrefutable conclusive prima screenshots indisputable disprove empirical provable validate incontrovertible purported plausible corroborating substantiates confirming confirms causal imperviable circumstantial scintilla corroborate alibi doubting certification indicating testimony testament disputing undeniable attested notarized attests refutation antidote convincing testify probable verified certify information trial presumed cast disproving voucher hint prototype disproved withstanding reasoning soreness nuisance pang sting trouble pressure painfulness tenderness cramp poison rack numbness neuralgia urodynia tightness causalgia myalgia meralgia nephralgia dysmenorrhea mastalgia cramping arthralgia feeling nagato keratalgia glossalgia morphine colic tension pleurodynia excruciate burn sensitivity irritation podalgia bleeding permanent spasm sadness shock photalgia odynophagia tingling weakness annoyance acute strain referred itch itching blood recovery stiffness frustration botheration bloating sedation insomnia healing poorly badly upset sickly nauseous worse disability disorder inauspicious ominous malaria dizzy viral indisposed nauseated seasick ailing afflicted convalescent hallucinating woozy addiction accident mononucleosis tuberculous bilious feverish personality hospitalization tubercular gouty hostile delirious spastic diabetic faint dyspeptic bedfast substance bronchitis unpropitious scrofulous giddy swooning consumptive gastroenteritis tonsillitis queasy peaked flu airsick meningitis eating livery neurodivergence surgery cold suicide carsick sneezy feverous learning ear diseased glandular staph unhealed psychological psychosis drug vertiginous instability strep flulike fungus parasite bacterium prion microbe organism toxin recombinant virulence fungal coli biology parasitic pest causative pathogenic insect fungi germ nematode spore pathogenicity trematode salmonella anthrax antibiotic saprophyte epidemic smallpox influenza antigen pandemic mycobacterium leprosy mycoplasma microbial pneumonia helminth escherichia chemical immunocompromised plasmid micro-organism bioweapon protein biohazard lactobacillus haploid mutation diploid commensal immunosuppression avian hookworm conspecific overgrowth bacillus flora genetic pathogenesis cholesterol endemic mycelium outbreak carcinogenic organismal bio pathology unicellular virology greek h5n1 zygote contagious pathologic serology yersinia ecological alga blight metastasis photosynthetic biologic carcinogen cytology susceptibility intercellular contamination virulent waterborne phytophthora pathological somatic tumor genetically extracellular malware infects undifferentiated intracellular polyploid mutated human microbiology mushroom malignancy precursor algae mammalian metabolism inoculation angiogenesis prokaryote protist eukaryote neurobiology chemotherapy tuberculosis gout eukaryotic virion consignment embryology dysplasia biological cellular histology coevolution filariasis morphogenesis cambium symbiotic multicellular chemotaxis parthenote chytridiomycosis biologist biotherapy vacuole listeria biologically penicillin toxicity infective myrmecophile bacteriology vesicle insulin h7 streptococcus bacteriophage phage protozoan regulome lectin pestis bovis aphid genotype mutagenesis biofilm infestation antibacterial lichen zoonotic antiviral genetics itis pseudomonas embryo maternal placenta gestational pregnancy foetal fetus newborn fertilization uterus womb umbilical hormone placental cervical reproductive cardiac hemoglobin embryonic perinatal stillbirth anatomy ductus prenatal preterm calcium weight phosphorus hypercorrection amniotic neural fertility heroin hormonal lanugo abnormal infertility neonatal ovulation deformity sac mother asphyxia circumcision respiration metabolic hypothermia ovarian embryonal abortion american larynx osteoporosis stem miscarriage circulatory induced mortality nursling breast vegetative conception obgyn sids english limb pediatric spinal fracture incidence estrogen ultrasound diabetes tumour ovary mouse neonate childhood malformation menopause neuronal cord urinary heartbeat apoptosis mitochondrial maturation childbirth sperm thinning neurologic bladder clot sterility implanted genital puberty cancerous colorectal bone utero viability imprinting gallbladder p53 prevents pediatrics babyish babe pediatrician babyhood creche folk infancy isidore polycythemia commonwealth hypocalcemia infantile mammogram intrauterine foundling gynecology unborn immature babysitter toddler anesthesia childish babysitting offspring dermatology orally biomedicine therapeutic reflex thought react backlash feedback resistance overreaction retort rejection comment criticism tropism riposte chemistry knee reacted kinesis catalyst elicited lukewarm widespread denunciation condemnation combat overreact commentary rebuke outcry dysfunction paresis palsy blindness deafness convulsion atrophy degeneration stun botulism hemorrhage complication amnesia cerebral stroke lucid paralyzed paralyzing poliomyelitis quadriplegia degenerative hemiplegia paraplegia comatose hallucination nightmare quadriplegic seizure paralyzes stalemate gangrene inaction helplessness delusion hypnagogic astral clumsiness strangulation irreversible flashburn disfigurement malaise electric rewind incurable coma inability sclerosis disfunction psychotic impotence function ed procrastination dyssynergia nephrology erectile surgical renal impairment cardiology bipolar schizophrenia epilepsy anemia adhd cardiovascular stigma gastrointestinal libido dysregulation dysautonomia pot pelvic arthritis corruption premature cpps dementia rheumatoid alzheimer memory sexual activation baggage adrenal cystic therapy bowel retardation tic systemic facial autonomic neurology sensory thyroid sepsis recurrent myeloma constipation degeneracy urology musculoskeletal phobia laryngeal ocd otology peptic autism vascular hematology connective hyperactivity congenital leukemia operation asthma restlessness anorexia osteoarthritis ophthalmology rheumatology radiotherapy nonspecific oncology anesthesiology gastroenterology progesterone diagnostic psychotherapy psychiatry palliative neurologist anticholinergic operable clinician anticoagulant functionality paregoric antihistamine apc placebo purgative immunotherapy specialist splint laxative

| |
|---|
| aspirin pasteur breakdown fog responsible chance associated inflicted scalable available priced draining rending facie vein arteriosus fluid tube development etymology venosus bud jerk sightedness |
| keywords for individual factor<br>individual personal private religion islam spiritual judaism christianity worship faith buddhism ritual secular clergy church hinduism sacred belief pious fundamentalist buddhist deity orthodox catholic interfaith churchly devout catholicism churchgoing god cult prayer muslim cultural atheism islamic christian sikhism ecclesiastical orthodoxjudaism ethnic symbol creationmyth evangelical protestant cleric dharma spirit votary churchlike scrupulous god-fearing cenobite confessional taoism religiousbelief moral theological religionist chapel conservative religiouspractice religiousperson agnosticism theology sunni mosque religio holy scripture culture vaishnavism priest religious jainism nicenecreed reformjudaism religiousbeliefs racial polytheism philosophical sikh human tradition sermon zoroastrianism religiousminorities romancatholic festival sacrifice trance banquet funeral traditional initiation meditation ideological matrimony mandaeism religiousaffiliation temple synagogue gnosticism monotheism religioustolerance religioussymbols catholicchurch bishop baptist easternorthodoxchurch presbyterian pope episcopal anglican liturgy easterncatholicchurches fullcommunion uniate romancatholicchurch pentecostal methodist holysee bishopofrome lutheran calvinist eucharist archbishop diocese jesuit cathedral doctrine anglicancommunion ethic pontiff greeklanguage rome latinliturgicalrites apostolicsuccession particularchurch mormon congregational episcopalian broad-minded nun orthodoxy melkite smyrna nationalist latinrite irish roman latitudinarian jewish protestantreformation ignatiusofantioch immaculateconception protestantism jesuschrist pentecost sacredheart parish christiandenomination immaculateheart cardinalnewman msgr jerusalem nationality ethnicity indigenous tribal minority pagan ancestor multicultural tribe ethnically native anthropology ethnical nationalism racist racism heathen separatist race folk sectarian ethnicminorities serb iraqi malay communal dialect meltingpot serbian gender contemporary immigrant political ethnictensions pashtun partisan fredrikbarth genocide culturaldiversity ethnicminority latino asian uyghur ethnicbackgrounds affirmativeaction ethnicstrife caste hmong diversity apostolic papal holiness pontifical apostle ecumenical priestly papacy liturgical apostolical nuncio vicariate benedictxvi vicar prefect popebenedict holyspirit episcopate encyclical gospel protonotary eparchy cardinalratzinger theotokos prophetic saint eastern blessedvirginmary communion monastic lordjesuschrist popebenedictxvi newtestament benedict holyghost oneness karolwojtyla franciscan doctrinal pontificalcouncil patristic orthodoxchurch anglicanism catechism usccb theologian discipleship incarnation faithfulness orthodoxchurches presidingbishop episcopacy sacrament diocesan ratzinger evangelization laity prelate pastoral sacramental eucharistic cardinal christ consecrated nunciature vatican titular exarchate patriarchal curia consecration exarch fourmarksofthechurch bishopric suffragan ordination patriarchate ordained synod priesthood chaplain pius patriarch coadjutor curate legate archbishopric infallibility congregation ascension enthroned consul prothonotary lds |

constitution polity plebeian xii constantinople exhortation xvi immaculate presiding conclave delegated canonized salesian seminary iau xiii magisterium ecclesial synodal apostolate conciliar ecclesiology presbyter ecumenism sacerdotal ecclesia mariology oblate christology benedictine chrysostom colossian thessalonian foundress monasticism augustinian synodical fulness cistercian sanctification canticle pelagius eusebius schismatics almsgiving diaconate prelature ecclesiastic areopagus ecclesiae norm rule role average standard criterion expectation touchstone normative southerncalifornia pattern guideline value knowledge experience information skill wisdom cognition expertise perception understanding learning awareness epistemology insight ability vocabulary erudition mind skepticism communication understand episteme motivation generalknowledge familiarity history knowhow encyclopedicknowledge philosophy subject domainexpertise interpersonalskills comprehension scientificmethod learned intuition plato education intellectualcuriosity reasoning collectivewisdom competence invaluable talent training versed interest truth impart gleaned intelligence knowing knowledgeability consent extensive resource nous lore opinion socialnorm conformity socialization socialcontrol socialpsychology socialinteraction collectivism deviant cheerleading bulimia sociality sociable party society fiesta multiparty jamboree festivity soiree kuomintang socialism partyness gregarious partygoer festive partymeister partyism nonparty bodylanguage shindig partyer orgy intraparty preparty fete housewarming attitude behavior mindset posture personality stance approach position mood mentality orientation tone tendency disposition demeanor viewpoint inclination decubitus ectopia bodilyproperty asana respect lotusposition unerect mentalattitude openmindedness mannerism singlemindedness work lackadaisical mentaltoughness killerinstinct easygoing ruthless hardnosed perspective outwardly cocky hardlinestance effort ego consciousness conscious unaware sensible mindful cognizant awake cognizance sensitive mindfulness know knowingness cognitive cognisant careful raiseawareness attention heightenawareness outreach educating heedful control acceptance alive awarenessmonth promote publicize positioning breastcancerawareness visibility redolent reminiscent healthy lifestyles informed sense psa identity demographic citizenship endogamy language acculturation ancestry irrespective stereotyping preference regardless hispanic background geography chauvinism discrimination affiliation intermarriage ethnogenesis mythology income salary profit wealth wage money revenue pension expense unearnedincome netincome unearnedrevenue net earnings yield return saving benefit cost debt taxable fund payroll expenditure payment employment insurance spending dividend cash account credit asset premium fee caput marginal investment purchasing gain minimum property loan unemployment portfolio job taxed costofgoodssold lucre employer planned employee workforce employ labor engagement occupation workplace workload piecework jobless housing utilisation utilization usage hiring worker employable booking exercise publicservice jobseekers gainfulemployment labour unemploymentrate nonfarmjobs gainfully employed contract underemployment gratuity financial nonfarm joblessness underemployed joblessrate career apprenticeship volunteer jobseeker unskilledworkers educationalattainment commission workforcedevelopment age old mature karma lifetime young epoch senesce oldness older oldage timeoflife antiquity maturate life geton eld majority historicperiod immature senility dotage newness mentalage adult ageofconsent child chronologicalage maturity ageofreason forty elderly youngest puberty antique midtwenties lifespan lifeexpectancy seniority ageism thirty adolescence mid midthirties twenty sex sexuality masculinity male femininity feminine genderidentity transgender sexual intersex grammatical female feminism woman equality sexology inequality homosexual genderrole genderstudies masculine hijra homosexuality androgynous androgyny epicene sexism johnmoney judithbutler genderqueer worldhealthorganization senior date heroic bold courageous courage bat gallant valiant desperate spirited resolute stupid warrior intrepid unafraid dauntless bravery audacious undaunted stalwart stouthearted valorous daring endure crazy selfless chrome gutsy strong resourceful heroically unfearing tenacious bravely bravesouls courageously braveout spunky legendary gamey smart foolish default brilliant tough adventure attack reckless insane ninja curious risky hearted colorful foolhardy mettlesome confident gritty braw ferocity eager gungho gallantly selflessly ultimatesacrifice gamy dust defy lucky braving lionhearted brave braver strongwilled nobly honest adventurer chromium grateful drunk duckduckgo venture dumb firefox edge valor proud braved ferocious persevere comfortable patriot skilled sojourner stunning uphold unsungheroes noble opera dpe fearlessly yuusha wise persevered inspiring lonely accessible accurate active adaptable admirable agreeable ambitious amiable amusing appreciative articulate artistic assertive athletic attentive attractive balanced benevolent big-thinking breezy calm capable captivating caring casual cautious charismatic charming cheerful clear-headed clever compassionate complex conscientious considerate consistent constant constructive contemplative convincing cooperative courteous crafty creative cultured cute decent decisive dedicated delicate dependable determined dignified directed disciplined discreet dutiful dynamic earnest educated efficient elegant eloquent empathetic energetic enthusiastic ethical exciting expedient experimental extraordinary extravagant fair faithful farsighted fashionable fiery flamboyant flexible focused folksy forgiving forthright frank freethinking friendly fun-loving generous gentle genuine glamorous good-natured gracious hardworking helpful honorable humble humorous hypnotic idealistic imaginative impressive incisive independent individualistic innovative insightful intelligent intense interesting intuitive inventive kind knowledgeable likable logical loquacious lovable loyal lyrical mellow methodical moderate modern modest neat objective optimistic orderly organized original passionate patriotic peaceful perceptive personable persuasive playful polished popular practical precise principled profound protective purposeful punctual rational realistic reflective relaxed reliable respectful responsible romantic self-sufficient sophisticated spontaneous stable steady studious sweet sympathetic thorough tidy tolerant trusting uncomplaining undogmatic vivacious well-rounded witty abrasive abrupt absentminded aggressive aimless aloof amoral angry antisocial anxious apathetic argumentative arrogant barbaric belligerent bewildered bigoted bizarre bland blunt boorish bossy braty brutal callous cantakerous careless catty childish clumsy cold complacent conceited conformist confrontational confused controlling cowardly crass critical crude cruel cynical deceitful deceptive defeated defensive

| |
|---|
| demanding destructive devious discontented discourteous dishonest disloyal disorganized disrespectful disruptive distant distractible disturbing dogmatic dominating domineering dull egotistical envious erratic escapist evasive fanatical fearful fickle finicky fixed flaky forecful forgetful frightening frivolous fussy gloomy gossipy greedy grumpy guarded gullible hateful helpless hostile humorless hypocritical ignorant impatient impractical impressionable impulsive inattentive inconsiderate indecisive indifferent indulgent inflexible inhibited insecure insensitive insincere intolerant irrational irresponsible irritable jealous judemental know-it-all lazy macho malicious manipulative materialistic meddlesome melodramatic messy mischievous miserable miserly misguided moody morbid nagging naive narcissistic narrow-minded needy negative neglectful nervous neurotic nosy obnoxious obsessive opinionated oversensitive panicky paranoid perfectionistic pessimistic petty picky pompous possessive power-hungry predatory prejudiced presumptuous pretentious prideful pushy quarrelsome reactive regretful repressed resentful rigid rude sad sadistic sanctimonious scary scatterbrained scheming secretive sedentary self-centered self-destructive self-indulgent self-loathing selfish shallow shortsighted sleazy sloppy sneaky spoiled stingy stubborn subservient sullen superficial suspicious tactless temperamental thoughtless timid unappreciative uncaring uncommunicative unconvincing uncooperative undisciplined unemotional unethical unfriendly ungrateful unimaginative unintelligent unprincipled unrealistic unreliable unruly untrusting uptight vain vindictive violent volatile vulgar vulnerable weak-willed whiny withdrawn workaholic worrier anticipative aspiring challenging companionly conciliatory debonair dramatic ebullient esthetic felicific firm forceful hearty high-minded incorruptible inoffensive insouciant invulnerable leader leisurely magnanimous many-sided meticulous multi-leveled observant painstaking perfectionist protean providential prudent responsive reverential rustic sage sane scholarly self-critical self-defacing self-denying self-reliant self-sufficent sentimental seraphic serious sexy sharing shrewd simple skillful sober sporting steadfast stoic suave subtle systematic tasteful teacherly tractable upright urbane venturesome well-bred well-read winning youthful artful ascetic authoritarian boyish businesslike busy cerebral chummy circumspect competitive confidential contradictory crisp dreamy driving droll dry earthy effeminate emotional enigmatic familial formal freewheeling frugal guileless high-spirited hurried iconoclastic idiosyncratic impassive impersonal invisible irreligious irreverent maternal moralistic mystical neutral noncommittal noncompetitive obedient old-fashioned ordinary outspoken paternalistic physical placid predictable preoccupied progressive pure questioning quiet reserved restrained retiring sarcastic self-conscious sensual skeptical smooth soft solemn solitary stern strict stylish subjective surprising unaggressive unambitious unceremonious unchanging undemanding unfathomable unhurried uninhibited unpatriotic unpredictable unsentimental whimsical agonizing arbitrary artificial asocial boisterous brittle calculating cantankerous charmless coarse colorless complaining compulsive condemnatory contemptible conventional criminal decadent dependent difficult disconcerting discouraging disobedient disorderly disputatious dissonant easily discouraged egocentric extreme faithless false fanciful fatalistic fawning fraudulent graceless grim haughty |

| |
|---|
| hedonistic hesitant hidebound high-handed imitative imprudent incurious inert insulting irascible mannerless mechanical melancholic mistaken money-minded muddle-headed narrow nihilistic obvious odd offhand one-dimensional one-sided opportunistic oppressed outrageous passive pedantic perverse plodding prim procrastinating provocative puritanical quirky reactionary regimental repentant ridiculous ritualistic ruined scornful slow sly small-thinking softheaded sordid steely stiff submissive superstitious tasteless tense thievish transparent treacherous trendy troublesome uncharitable uncreative uncritical unctuous unhealthy unimpressive unlovable unpolished unreflective unrestrained unstable vacuous vague venomous weak willful allocentric herioc leaderly liberal manly maticulous nonauthoritarian planful pruposeful unfoolable crebral huried old-fashined stoiid unpredicatable unreligious airy arrogantt astigmatic biosterous complaintive dirty dissolute enervated excitable grand libidinous mannered mawkish mealymouthed meretricious monstrous natty negativistic overimaginative pharissical phlegmatic profligate pugnacious rowdy shy silly single-minded strong-willed unself-critical venal well-meaning wishful zany amenable approachable averse biased candid commanding condescending contemptuous dark defeatist diligent disengaged disgusted dismissive enterprising impartial impolite inhospitable joyous lighthearted overconfident patronizing pleasant poor reasonable reluctant resilient somber thoughtful tireless unapproachable unbiased unprofessional unwavering willing animated authoritative bad buoyant carefree committed dejected detached devoted disagreeable disgruntled disinterested distrustful engaged fragile giving glum grounded hopeful interested lively motivated outgoing polite positive professional remote sincere snobby unaffected unassuming unenthusiastic unpretentious unwelcoming accepting adventurous affectionate ambivalent angsty annoyed antagonistic avoidant awestruck broadminded cheerless closed-minded cold/cold-hearted concerned conciliat contrarian depressed diplomatic disdainful expectant flippant forgiveness fretful frustrated furtive hopeless imperious imperturbable indignant inquisitive intransigent joyful/joyous judgmental loving malevolent mean-spirited morose no-nonsense obsequious open parsimonious patient paternal pragmatic querulous recalcitrant resistant reticent reverent sardonic self-assured self-effacing self-righteous sorrowful spiteful traumatized vigilant warm freedom loyalty creativity humanity invention generosity integrity finesse openness order advancement joy excitement goodness involvement beauty honesty kindness excellence innovation quality commonality contributing spiritualism strength entertain affection cooperation encouragement pride clarity charisma hum leadership renewal betrue contentment friendship balance compassion fitness professionalism relationship patience prosperity wellness gratitude grace endurance facilitation effectiveness fame justice appreciation willingness trustingyourgut self-respect abundance reciprocity enjoyment entrepreneurial happiness harmony worried despairing toothers maternalistic open-minded bitter sharp snobbish tim indefatigable unsociable immovable hypervigilant explosive |
| keywords for interpersonal factor<br>family household friend house parent son uncle home mother nuclear grandmother father marriage sister grandfather child cousin relative nephew kin brother extended clan ancestor |

grandson aunt niece sibling daughter immediate grandparent loved stepsister stepfather mom couple tribe wife fiancée husband lineage folk subfamily dad granddaughter stepson stepmother sib eldest spouse individual love unfathered kinship conjugal grandma kid personal stepbrother stepdaughter younger partner relationship latin language consanguinity kinsperson grandaunt maternal daddy fiancé grandpa parental neighbor stepdad paternal yourself remarried stepchild baby grandchild mob human anthropologist friendship someone boyfriend mafia loving kitchen papa pa male forefather pater founder dada sire beginner priest father-in-law padre hypostasis beget begetter engender generate father-god elder church mentor bring forth don granddad twin godson infant toddler orphan childhood teen boy person girl offspring minor adult youngster preschooler tyke babe juvenile adolescent imp kindergarten newborn kindergartner kiddy puberty teenager urchin bairn waif ren daycare foster firstborn rapist woman bambino trafficking endangerment slavery abortion scallywag immigrant aid babyless kinderwhore secondborn school kitten sextuplet attention deficit hyperactivity disorder laundering prodigy slave pickaninny fetus sprog progeny alimony maternity parenting motherhood parenthood guardian adoption spousal prenatal pregnancy motherly familial marital fatherless fatherly autistic custodial stepparent daughterly filial involvement rearing societal explicit statutory guardianship homeschooling mat premarital unwed adoptive custody homeschool truancy babysitter teacher adolescence class caregiver nanny litem dcfs social caretaker mothering stepfamily cultural peer permanency older dyfs caregiving childcare legal medical adulthood paternity childrens sitter community cyf support childbirth emotional inheritance pre natal reproductive neighborhood society village sidebar group local public communal neighbourhood congregation area parish organization ecology convent inca town close communitywide fandom grassroots volunteerism catholic identity volunteer residential betterment environment subreddit tightly server ethnography guild revitalization volunteering philanthropy forum team nonprofit interfaith institution classroom student college education university educate library primary middle elementary schoolroom private grade blackboard work schoolhouse sport schoolmate classmate gymnasium schoolteacher schoolchild teach scholastic graduated varsity secondary prep cultivate academy campus schoolers online coeducation schoolgoing homeroom career residency grader vocational schooling conservatory tuition preschool living gym playground coaching stanford chalkboard minischool cyberschool schooly tutor tutorial practice serf special acquaintance roommate freshman schoolfellow colleague valedictorian schooler sophomore girlfriend graduate fellow schoolboy fiance upperclassman coworker headmaster guidance counselor collegian classman co-worker fifth assistant principal schoolfriend buddy ex brianna kayla sweetheart teenage homecoming queen carly emilee teammate kaitlyn backcourt mate arielle kaleigh pal grad classer alumnus ally comrade associate champion admirer companion chum flatmate lover quaker supporter confederate amigo mortal befriended somebody befriend crony member protagonist dear dodo homeboy crush roomie guy intimate employee employer job democracy company salary gratuity workmate piecework commission stock worker clerk workplace party pencil pusher corporation housing collaborate workforce tenure contract workfellow housewifery toiler overwork wage workpiece toil employment freelancer pairwork labour workable not-for-profit teamwork ironwork bos rework employ socialism erg labourism fink freelance jobless coursework socioeconomics workling pejorative collaborator interwork labwork manager benchwork weisure swink yakka allwork gridwork cicero insurance workstead ssp prework taskwork supervisor workshed paperwork bushwork lacework leatherwork workgroup underwork customer clerical workful stuccowork spadework nonworking taskmaster workfree cobwork metalwork sourcework inwork brassworks trip travel racism collegiate academic seminary educational undergraduate academia dartmouth professor lecturer uni faculty graduation bachelor polytechnic academe semester postsecondary ucla ivy league extracurricular highschool canadian imprs matriculation undergrad academically berklee graduating tertiary teaching ncaa occupational studio discrimination disability ergonomics workshop bullying hygiene profession home-office roundhouse waterworks laundry shipyard bakery bakehouse brokerage beehive tannery pit colliery business lab farm creamery laboratory shop fishery forge location gasworks smithy glasswork professional lumberyard piscary bakeshop ropewalk hiring management economic harassment mental environmental union yale institute sorbonne mit princeton columbia penn oxford bologna multiversity cambridge scholasticism paris univeristy emeritus phd aristotle urbana adjunct uc san diego doctorate ph.d massachusetts amherst tuft honorary dean monash oberlin engineering collage distinguished carnegie mellon astrazeneca studentship scholar study tenured emory course thesis mechanical zoology neighboring border lebanon object abut butt adjoin march edge inhabit dwell populate live neighbour street neighbouring landlocked gardening impoverished bordering wedged destabilize foe caucasus restive yemen tajikistan syria balkan neighbourly noise sudan erstwhile archenemy wracked doorstep armenia macedonia landlord troubled albania rift northern bunk next-door accuses ethiopia embattled russia owner attache estranged nearby eritrea rival tujue restaurant arround mon old major elderly chief seniority aged fourth-year soph captain redshirt letterwinner adviser superior guard letterman higher-ranking precedential stater lead majoring standout vice president kelsey external underclassman staff executive tayler leading rebounder representative devan libero experienced signee neuqua valley regional intermediate setter returnees requesting anonymity orally committed recruited director redshirted disabled returning starter eiu elyse leadership scorer medior honor recruit veteran blocker kaila homewood flossmoor lyndsay architect managerial swingman upperclassmen attending pleno pwd doyen octogenarian oldish engineer leader centenarian principle skilled geriatric olden retired ancient antediluvian retirement exec scientist antiquarian eld assisted anciently antiquary nursing important established immemorial managing specialist accountant antiquity paleo regular bar cafe cafeteria bistro diner eatery buffet cuisine sushi deli hotel dinner waiter steakhouse grill brasserie menu coffee dining pub canteen waitress tavern greasy spoon tearoom chef pizzeria lunchroom brewpub breakfast store coffeehouse grillroom tapa fastfood parlor rotisserie retail chophouse grille chang teashop inn restaurateur establishment nightclub wok donut garden bennigan supermarket catering meal tgi friday mall brewery pompeii salon grocery service applebee smokehouse hospitality cantina club lobster shopping outback barbeque melting pot delivery takeout bq takeaway quiznos banquet hall café lounge building venue edifice teahouse entertainment take-out movie greece vegetarianism rome station commercial market gourmet upscale boutique chain motel cooking manhattan snack tastet apartment arcade posh theater thermopolium convenience cinema warehouse truck winery nightlife driver trendy midtown cocktail stall barbecue ritz museum crowded music bookstore cottage homemade app bartender bartending resort butcher casino parisian proprietor garage lodge merchant franchise lobby she your herself honestly aquarius capricorn goat archer sagittarius scorpio scorpion balance libra virgin virgo crab leo taurus gemini ram bull jew aries slav gentile amerindian negroid caucasian negro asthmatic african traveller unfortunate sensualist traveler precursor percipient forerunner beholder compeer themselves intellectual intellect aborigine inhabitant aboriginal stranger dweller gypsy individualist gipsy entertainer therapist experimenter technologist others disputant automaton contestant coward bot commoner himself modifier captor appointee appointment own chassis bod amateur mommy mum pappa hey sissy abby auntie granny stepmom gran sweetie timmy mummy bro ma snr step mama grandad me abe momma coach fella fatherese fatha daddio missus fatherdom merfather dude biofather pun telegony stepuncle grandniece da matrist hubby adjective agent bear fatherling probole boomer stepaunt dom king sperm fathership grand frater belly dog crazy cop remember brotherly gramps triune mam stepmum jock hygiea kinsman trainer grampa chubby donor parentless santa corny cub stud kinswoman chub our grandkids stepnephew progenitor poppa anymore fra hate pappy book writer artist poet post novelist essayist coauthor biographer playwright bronte journalist burroughs generator historian authoress novel publisher content researcher story illustrator scriptwriter paragrapher sociologist speechwriter source treatise pamphleteer title poem fiction literary screenwriter psychologist editor biography bestselling literature authored space coauthored columnist review series narrator evolutionary biologist syndicated cookbook reader speaker authorship mangaka complete guide artemus ward memoir philosopher italo calvino kate chopin contributing article contributor penned investigative translator andersen ghostwriter burnett browne burgess nonfiction history pulitzer bestseller developer designer maintainer dummy actor audience user malcolm gladwell blogger encyclopedia youtuber myth autobiography publishing activist fic ph.d. filmmaker composer dev poster reporter manga podcaster wolfe mann flair autograph poe comic cartoonist harris published host interviewer anime lawyer diary manuscript dodgson seller sponsor singer producer narrative presenter medium maker librettist polemic lyricist versifier critic storyteller thinker podcast musician photographer plot influencer comedian rhymer shaper commentator tragedian carroll aiken alger redactor inventor drawer guru chandler anderson algren asch aragon breastfeeding immature neonate foundling nursling puppy spoil coddle womb unborn godchild pamper mammal indulge cocker featherbed cosset mollycoddle quadruplet papoose infantile birthing pup diaper premature pacifier uterus expectant jayden pet pushchair internship hire money vocation duty subcontract workload position stint caper speculate chore menial thankless overqualified fulltime severance package income assignment role occupation résumé waitressing secretarial résumés rehiring vacancy seeker hobby resumé field scut gainfully

| |
|---|
| employed intern apprenticeship rehired laborer mother-in-law brother-in-law sister-in-lawson-in-law daughter-in-law widow widower spinster account accounting adjustment admiralty alcohol athletes athletic atm atmospheric audio-visual audiovisual automobile automotive avionics baker ballistics bus cabinet camp |
| keywords for system factor<br>system policy law administration legislation political insurance stance government principle governance strategy rule diplomacy agenda plan guideline reform priority proposal legal initiative foreign politics eligibility procedure policymaking regulatory management directive constructive interventionism privacy mandate regulation election ballot referendum poll veto elect voting electoral voter majority suffrage balloting turnout franchise plebiscite secret voted vote electorate overwhelmingly state approval senate endorse cast debate revote parliamentary nominate democracy nomination supermajority casting elector reelected ratify support precinct amendment elected constituency abstention absentee provisional biden scoop trump outflank ruff crossruff trounce gop trumping trumped reign trample overridden negate republican undercut russia usurp overwhelm outlast supersedes stymie maga eviscerate politician diplomatic polity diplomatical republic public aristotle partisanship economics politically partisan politicking politic suave religion politico police nationalism regionalism war ideology monarchy activism expedient geopolitics parliament democratic senator lawmaker candidate governor congressman mccain liberal incumbent democrat lieberman pelosi congressional crist conservative dem huckabee lieutenant gubernatorial hillary officeholder dems gopers libertarian minority councilman reps left superdelegate leftist shay progressive fiscally alderman pol representative president dick hospital clinic doctor patient nurse medical surgery ambulance surgeon icu outpatient sanatorium physician intensive infirmary psychiatric treatment care hospice healthcare medicine nursing morgue radiology inpatient paramedic trauma dentist emergency lazaretto antihistamine nanomedicine teaching memorial maternity mental quarantine methodist christiana hospitalized dermatologist hospitalize psychiatrist ophthalmologist multidoctor pediatric rehab psychiatry regional vet doc md therapist gastroenterologist veterinarian neurologist pediatrician medico allergist gp harvey ross gilbert manson medication gregory dr. gynecologist specialist oncologist cardiologist internist medic pharmacist obstetrician radiologist avicenna scientist schweitzer hakim hodgkin lawyer midwife veterinary clinician checkup abortionist psychologist student general professional profession practitioner professed collegiate athlete expert trained professionalism qualified amateur technical pro journeyman nonrecreational nationally semiprofessional organisation presidency organization federal bureaucracy establishment disposal aide authority campaign executive anarchy hierarchy brass judiciary congress adminstration sedation commission predecessor clinical pharmacy lab physiotherapy dispensary therapy center pediatrics counseling centre daycare orphanage laboratory urologist dental ministry confirmed volunteer civil bureaucrat secretary unofficial officer functionary authorized spokesman authorised formal authoritative umpire diplomat referee established anonymity semiofficial statement bureau |

| |
|---|
| deputy prescribed announcement military legislature country governing privatization coalition civilization aristocracy constitution opposition anarchism cabinet governmental sovereignty patrol sheriff constable arrest constabulary policeman gendarmerie france crime trooper civilian paramilitary security enforcement detective riot investigator guard bow army plainclothes assailant justice arrested jurisdiction fbi prosecutor front-line republication obama warfare economic protest entity manufacturer advisory committee pharmaceutical drug prescription anticholinergic biopharmaceutical nsaid pharmaceutic insulin biotech antibiotic aspirin antidiabetic pharmacology benzodiazepine therapeutic immunosuppressant biotechnology biomedical analgesic corticosteroid antifungal medicament preventive antiemetic anticonvulsant antidepressant substance antitussive decongestant bronchodilator antidiarrheal antispasmodic drugmaker ppe universal chiropractic oecd information ecosystem charity dietetics midwifery optometry organizational medicare deductible sanitation systematic organon allied illness dentistry diagnosis psychology medicaid provider telecommunication pharma drugstore store apothecary oncology anesthesiology dermatology gynecology walgreens chemist neurology nutrition urology chemists apothecarys cosmetic grocer pbm walgreen caremark rx prescriber hba pbms minuteclinic bookstore greek med gun grocery obstetrics ob medco medicate immunology gastroenterology alternative splint placebo apc remedy biology anticoagulant acupuncture pill ophthalmology shot paregoric diet zyloprim minister ciziten indemnity grant hoax pastor rector ministerial curate priest clergyman parson prime minster ambassador interior defence leader mp premier preacher permanent chancellor mla chief parliamentarian innen advisor adviser labour authorization mandatory lockdown prescribe delegate authorisation fiat passport depute designate dictate force mandating constitutionally statute dominion moonstone mask legislated injunction encourage pledge territory recommendation restriction enforce requirement commandment prerogative statutory ban raison provision recommend prohibition rylais behest regulate rylai carte legislate coerce enforceable exemption request penalty reauthorized writ dispensation shurelyas testing vaccination pas territorial certificate license certification registration credential permit accreditation badge authentication ged certified receipt citation assurance designation licensing licensure licence scrip graduation baccalaureate photocopy minor attesting cert proof recertification notarized accredited bsc certificated training documentation medallion commemorative limber graceful sinuous pliable lissom lissome lithesome supply shortage silky scarcity supplied supplier nimble stockpile offtake velvety price availability shipment springy rubbery electricity outstrip quantity oversupply textured inventory buttery storage sinewy sensuous outstripping contoured tannic sonorous distribution lustrous sprightly procure satiny astringent punchy wholesaler balletic export logistics rationing nubby deliver potable capacity flow rationed purchasing insatiable authorize sanction release amend endorsed disapprove ratification ratified rescind authorise agreement unanimously introduce authorizing nix revoke override authorizes overrule decline solicit resubmitted opposed finalizes disprove conditionally opposes convene defer reconvene sticky reinstate overturn allocate tabled approved verify resolution adjourn disapproved cdc fda hhs atlanta epidemiology h1n1 who influenza epidemiologist nci sars |

| |
|---|
| prevention nih idc epidemiological immunization aid swine hepatitis researcher tuberculosis malaria fao recommand suggest advise urge commend advocate modify examine instruct advisable province commonwealth nation city status relationship merchantability provincial national loan concession permission allot allowance subsidy cede grantee grant-in-aid vouchsafe donation pension subsidisation lee gov resident nationality citizenship immigrant citizenry soldier servant activist jurist naturalized compatriot noncitizen ethnicity illegal participatory civic consul consulate expatriate consumer patriot tax seiko squadron jew lawfully announce denote statistic assessment investigation downvote composition briefing presentation recital exposé communiqué released court financial lawful judicial legitimate legality precedent litigation canon solicitor criminal jural constitutional licit valid barrister sharia parent compliance natural antitrust contractual legislative illegality liability tolerance supreme opportunism expediency district caucus nominee delegation gephardt samaritan george york molecule relaxant script co. gyn specialty service pranab reshuffle whip datuk mandated blanche plaque disruption stamped stamp citizen visa legend testament mukherjee chidambaram affaire seri etre hammurabi rules guidelines priorities initiatives approach foreign policy insurance policy zero tolerance policy regulatory framework stances directives constructive engagement party line refund political science laws strategies zero tolerance privacy policy agendas mandates community doctrines policies neoconservative process guiding principles currency peg decision contract action administrations interventionist research regulatory frameworks program principles practice accord pre emption ideological unilateral culture protocol dress codes regime platform treaty doctrine common minimum programme cato institute actions terms attitude hawkish international piecemeal approach procedures finance company policy studies regulations rhetoric organizational structure guidance advocacy proposals reforms leadership politicians science deliberation issues plank warranty position planning handbook admin floater preventive medicine surgical aesculapian scientific disease physicians conditions medicine man health care surgeons psychological nonmedical medical record examination psychotherapy biological religious medical exam humorism medical examination medical checkup health check alternative medicine mental health premedical hospitals cardiology medically nurse practitioner heath care disability emergency medicine tongue depressor nephrology podiatry physio antibacterial scrutiny naturopathic internal medicine recreational personal urological psych orthopedics outpatient clinics biomedical research physical therapy medical device kidney dialysis palliative care telemedicine nurses safety critically ill rec clinicians pharmacological genetic medics occupational tech bio ob gyn traditional medicine chemical developing country western world diagnostic industrial cardiologists hormonal spinal surgery surgical procedures technological physical examination nutritional interventional cardiology orthopedic surgeons differential diagnosis anatomical diagnostic tests anesthesiologist neurological coroner medical laboratory |
| keywords for communication factor |

communication language interaction social contact message telecommunication communicating messaging sign information transmission coordination communicator voice contact understanding intercommunication communicate wireless relationship conversation marketing networking collaboration trust nonverbal communication paralanguage miscommunication connection telephony speech writing connection connectivity connectedness dialogue medium teamwork service people eye contact chat texting communication visual communication customer exhortation feedback publication system talk to person share knowledge personality respect contagion telephone support transparency television movie film broadcasting video tv broadcast television set entertainment cable telly radio cable television cathode ray tube movie television receiver boob tube tv set telecasting bbc idiot box goggle box television program television system high-definition television cinema telecast channel audio advertising news internet picture medium satellite television kinescope dvd cbs itv analog television footage streaming simulcast comedy lcd television network nbc nipkow disk film serial john logie baird paris emi tuner interlacing selenium rca sound televise monitor general electric terrestrial television internet television sitcom plasma display computer cold cathode flat panel display show ntsc social color television youtube monochrome couch potato federal communication commission vhs television broadcasting corporation music broadcast online broadcasting system tvb satellite tv # # black-and-white aptn motion picture netflix video image audio picture television videotape tv tape recording dvd videocassette recorder documentary footage cassette picture camera multimedia telecasting image film videocassette clip movie screenshots youtube podcast technology demo animation photo broadcasting photo camcorder playback medium broadcast ntsc digital video visual communication digital blu-ray disc television receiver soundtrack video video camera studio television set highlight pic online stream youtube video internet screenshot videotape audiotape videotaped kinescope youtube.com grainy slideshow graphic gif hi def video recorder web cam content unedited videodisc high definition hd movie filmstrip web showing audio tape high definition song pic polaroid voice audio visual phone videogames mechanical television mtv cathode ray tube article computer projector jumbotron prerecorded post viewing camera music instructional video newspaper magazine paper news editorial newsprint gazette publisher daily article tabloid reporter journal columnist journalist headline medium publication obituary journalism column editor comic strip press advertisement periodical daily rotogravure publishing publication newspaper publisher newspaperman broadsheet newspaper pravda website daily mail production paywall news article cardboard daily news newsstand website app blog site webpage portal homepage internet intranet database world wide web computer email online television site document information wikipedia web server web page software url web website youtube web site page network hyperlink directory facebook twitter myspace html bulletin podcast official newsletter download list domain name video tv on-line publishing web browser extranet google polo play hockey blog perl page microsoft word processor netcraft xhtml server multimedia medium subscription chatroom service magazine internet site video newspaper social publication cern newspaper store anonymous etsy company forum unofficial news webpage program account released channel broadcast radio link press publication weekly application source platform distributed social medium social network twitter facebook network world wide web linkedin google viral marketing google+ event trayvon martin website telegraph reach blog telephone arab spring internet sociality social intranet technoself business cyber activist sociable socially microblogging pinterest cybernetic tumblr youtube extranet cyberspace cybernetics telnet computer-mediated communication modem virtual community informatics multiparty party predictor cyberinformation e-commerce virtual network fiesta cybertechnology cybersociology telecommuter cyberjunkie login festivity user jamboree teletype cybernetwork cybersavvy society newsgroup data kuomintang cyberterrorism mainframe sneakernet cyberjargon social network cyberpsychology cyberphilosophy cyberinteraction cybersuicide webcam computer professional network service netzine enterprise social networking multinetworked multinetwork soiree processor twitter facebook youtube reddit blog flickr microblogging jack dorsey biz stone tweet blogging chirrup wikileaks podcast sm iphone hashtag chat myspace internet instagram gossip character short message service barack obama dick costolo noah glass insta san francisco smartphone ashton kutcher mimicry chirp blog email google chitter website tumblr twitter tweeting twitter.com yahoo micro blogging site microblogging site twittering facebook.com micro blogging discord www.twitter.com tiktok perez hilton mashable hashtag # ustream myspace page social ipad blogosphere social networking service application software compete.com facebook page social networking site starling evan williams sb nation web gmail short code pixiv danah boyd twitch weblogs medium trending topic  homepage time skype bleacher report castle in the sky telegram friendfeed wikipedia blogged blogger yahoo.com the texted friendster popeater  tik bit.ly twitterer other venture capital here rt @ news onlyfans social networking website linkedin elon gawker sub online musk the new york time digg blog posting mtv.com patreon facebook connect tweet deviantart digital spy subreddit tweeter wordpress tok information data knowledge news evidence content advice database info detail intelligence report update document message entropy know code documentation material propaganda communication secret fact descriptor computer medium metadata pas datum input misinformation internet communication experience perception effort resource claim report resource contact context ice education reporting document informatics fact sequence help stimulation experience research factoid opinion example instance information theory readout tip predictor picture source selective information update learn idea explanation acquaintance link cognition information visit www.verizon.com insight discussion telephone internet call power phone telephony dial telecommunication telegraph landline handset cell phone online cellphone video ring call up speakerphone voicemail rotary dial cable call radio communication call in radiotelephone tele electric mobile phone email mobile alexander graham bell wireless ringing electrical dial tone on hook callback telecommunicate sound telephone receiver modem zoom answering machine smartphone mail dial telephone phone call data caller telephone call address whisper switchboard push-button telephone telephone system cellular dial phone computer virtual chinese broadband videophone light tree voice mail telephone cell verbal order voice dsl sm caller landline

| |
|---|
| telephone hang on tree fiber telephone set call waiting landline phone landline electricity hydro cordless phone phoning text message communication message communication chat statement communicate mail email courier letter telegram signal signal call reminder messenger theme dedication word information text warning post missive address exhortation substance content speech note pas remark subject matter direction pas on put across meaning view pas along comment mailer inscription cipher word voice sign message talk idea impression congratulatory message reference reply news e mail telegraphy missive send add ignore convey clarion call correspondence story text message stern warning greeting conversation transmission messaged strongly worded letter warning phone handwritten note statment image exhorting broadcast radio television cable air telecast rebroadcast simulcast circulate spread newscast transmit aired programme satellite publicize streaming multicast rerun cable television podcast pb stream broadcasting network televise broadcaster video show program television program audience televised publicise sow streamed news airing bare telegraph air go around print production send coaxial cable distribute programming medium disseminate live propagate disperse rtx beam broadcast medium publish film diffuse circularize record sportscast game show live television published studio voice broadcasted circularise recording commentary distribution broadcast bbc release talk show pas around broadcaster game professional journalism radio broadcast replayed share camera recorded telecasting channel televised nationally advertise receive social radio station announcer nationally televised stadium commercial cinema dvd simulcast ch . airtime eng post unicast article primetime fsn audio subnet graphic programing espn2 series simulcasting sound cstv distributed interview publication shared rumor gossip leak speculation rumour hearsay misinformation propaganda scuttlebutt sociology rumour psychology allegation accusation insinuation innuendo news disinformation bruit rumor circulating chatter rumor rumor mill rumor swirling suspicion factoid rumor circulated conspiracy theory myth rumor mongering unconfirmed report rumored gossip unconfirmed theory conspiracy theory joke hysteria rumor swirled completely untrue hoax controversy story tattle hype totally untrue grumbling unnamed source conspiracy theorist hyperbole announcement sell rant myth apocryphal digitimes frenzy report anecdote conspiracy categorically deny blogosphere utter nonsense revelation lie talk innuendo word untruth rumor fud idea legend urban sneaking suspicion meme categorically denied leak misconception national enquirer info fact speculation speculating categorically unsubstantiated assertion belief comment untrue assumption accusation guess insinuation information allegation fan falsehood informant FALSE reportedly narrative scandal inform misconception myth stereotype misunderstanding ignorance misperception misperceptions misinformation myth misinterpretation misunderstanding fallacy delusion illusion mistake error misapprehension question misconception idea stigma fallacy mistake lie shortcoming common misconception assumption implication confusion misnomer preconception negative stereotype perception dichotomy paradox drawback preconceived notion bias truism contradiction supposition disservice error issue misgiving stigma attached confusion stigma irony FALSE generalization faq prejudice falsehood gripe stigma associated rumor |

notion miscommunication pre knowledge inaccuracy ambiguity concern unanswered question misrepresentation miscommunications pitfall doubt misinterpretation opinion fear idea difference problem obstacle taboo stumbling block demystify theory negative connotation bias distortion preconceived idea educate criticism misinformed negativity gripe gender stereotype barrier worry mindset speculation attitude medium news internet broadcasting radio network youtube multimedia social website computer sound video television government telecast tv recording entertainment videocassette recorder fan dvd podcast audio cable television public telegraph online televise show footage people political clip digital documentary coverage aired television set politician kinescope press text television receiver art tech reddit message programming cable game marketing culture the mtv viewing society filmed telephone propaganda fan phone show linux content music cbc film gaming blog airwave movie video twitter tv set series game politics featured dating advertising hollywood released format interview big release information govt journalism simultaneously appeared left gov mockumentary book pop telly journalist medium carry streaming police telnet fandub newsgroup porn webcam electronics communication radiogram cultural influencer democrat technology dems telescreen movie mass medium internet book television radio medium information newspaper website blog magazine music news technology video game email animation press podcast gramophone record web site mass phonograph group film journalism billboard comic blimp mmorpg pamphlet mobile phone podcasts scheduling skywriting requiem subscription frequency organisation advertising journalist runescape multiplexing heap throng crowder crowd camera recording pay-per-view amass art multitude dubbing mass communication subtitle mob thring wireless batch ruck huddle computer mobile web ton congestion pile button social medium webcasting swad qr code high mass kilogram augmented reality podshow congregation agglomerate teem printing bulk calculator collective cell phone eucharist sacrament lot massif computer game sound electric conspiracy internet computer network social cyberspace network world wide web instant messaging google youtube facebook phone intranet cable online computer website web browser usenet social networking email web broadband ip address information data internet engineering task force multimedia app website cyber medium blogging database wireless modem communication electronic computer hyperlink reddit digital myspace chatroom cell link webcam microsoft wifi youtube.com browser gamer twitter virtual internet telephony news power newsgroup connection game peer-to-peer internet protocol suite hypertext transfer protocol laptop cybersociology supercomputer file sharing chat advertising search engine hypertext web site offline event festival story occurrence venue tournament circumstance phenomenon miracle occasion gala party celebration shiny occurrent case event materialization fundraiser quest reverberation expo fund raiser festivity wedding invitational concert issue spring fling bos holiday chili cook off banner fest game outcome person gala dinner time walk thon activity winterfest race pow wow day walkathon show season grand finale action incident oktoberfest place induction ceremony cookie banquet premium thing update mission firework display social event character situation campaign crusade story effort push election war battle run fight political campaign safari expedition race movement advertise electioneering campaigning blitz candidacy drive lobbying initiative agitate promote cause hunting expedition advertize take the field adventure military campaign stump reform campaign operation game election smear campaign action setting publicity advertise push publicize promote fight crusade campaign agitate communication press propaganda announce headline promotion propagandise bill poster ballyhoo edward bernays publicity tout proclaim marketing publicise service product persuasion sponsor praise publicis denote plug pompeii papyrus non-commercial jinan profit quackery omnicom interpublic propagandize havas bulletin sublimation ad advert mtv advertisement promulgate qvc news announcement notify coupon notification google tram newsagent television advertisement old medium outdoor advertising radio advertisement facebook direct mail new medium lobby marketing sale business commerce advertising charity marketplace mercantilism commercialism grocery product trade grocery store mart black market industry commercialize commodity branding retail food market economy agora market place sell merchandise promotion buyer "s" market monopsony bazaar oligopoly bull market labor market stock market seller "s" market money market grey market medium supermarket bear market social shop promotional wholesale mailing commercial stock exchange design brand greengrocery communication mercantile establishment hype management development merchandising marketing buyback marketer copywriting selling experiential marketing promotion marketing advancement personal publicity promo promote advertising relegation advertisement sponsorship encouragement advertise promoted hype furtherance promoting advert forwarding sweepstakes promotion promotional material promotional offs merchandising bonus microsite avoiding relegation prize demotion discount coupon spam raise morale boosting coca cola championship prize giveaway campaign demoted knockout coca cola sponsorship advertisement qualification relaunch boost mk don reward relegated crewe alexandra intertoto cup coupon ticket improve fa trophy goodie bag tranmere rover digital analog electronic virtual physical computer analogue electronics online interactive multimedia computing wireless audio mobile optical portable computational broadband data numeric computerized digitally digital imaging digital designer dtv number high definition digit dvd recorder streaming numerical digital camera traditional paper digital medium medium consumer electronics hi def vcr audio visual 3d social camcorder high definition hd mp3s crypto multiplatform disc iptv encoding print tech video recorder internet vod 3dtv numerator puter hypercomplex number archiving blu ray disc digital signage film google android youtube gmail search engine internet facebook aol adwords microsoft adsense apple mountain view
